# Effective screening strategies for safe opening of universities under Omicron and Delta variants of COVID-19

**DOI:** 10.1101/2022.05.04.22274667

**Authors:** Marie Jeanne Rabil, Sait Tunc, Douglas R. Bish, Ebru K. Bish

## Abstract

As new COVID-19 variants emerge, and disease and population characteristics change, screening strategies may also need to change. We develop a decision-making model that can assist a college to determine an optimal screening strategy based on their characteristics and resources, considering COVID-19 infections/hospitalizations/deaths; peak daily hospitalizations; and the tests required. We also use this tool to generate screening guidelines for the safe opening of college campuses. Our compartmental model simulates disease spread on a hypothetical college campus under co-circulating variants with different disease dynamics, considering: (i) the heterogeneity in disease transmission and outcomes for faculty/staff and students based on vaccination status and level of natural immunity; and (ii) variant- and dose-dependent vaccine efficacy. Using the Spring 2022 academic semester as a case study, we study routine screening strategies, and find that screening the faculty/staff less frequently than the students, and/or the boosted and vaccinated less frequently than the unvaccinated, may avert a higher number of infections per test, compared to universal screening of the entire population at a common frequency. We also discuss key policy issues, including the need to revisit the mitigation objective over time, effective strategies that are informed by booster coverage, and if and when screening alone can compensate for low booster coverage.

## Introduction

Almost three years into the pandemic, and, COVID-19, through emerging variants, continues to pose a threat to the in-person education in academic institutions. Over and over again, universities and colleges have faced an abundance of COVID-19 cases on their campuses [1], and have found themselves in a position to reformulate/reoptimize their infection mitigation strategies (e.g., vaccination mandates, routine screening, face masking, social distancing and hybrid learning practices), to adapt to the characteristics of the new Delta and Omicron variants, with an eye on future variants yet to come. A continuous reformulation of infection mitigation strategies will remain essential as COVID-19 continues to evolve with new virus variants, and as the current interventions (vaccines, testing kits, masks), their availability, and efficacy continue to change. With COVID-19 vaccination and testing kits already developed, the current challenges for effective mitigation differ from those faced at the beginning of the pandemic. Now the mitigation strategies need to account for new virus variants with different transmission and disease dynamics [2, 3], and vaccine effectiveness that is not only imperfect and decaying over time [4], but is also variant-dependent.

Indeed, as the year 2021 came to an end, the then-dominating Delta variant of the virus started to be replaced with the emerging Omicron variant, which, by early January of 2022, has become the primary variant that was causing almost all COVID-19 infections in the United States (U.S.) [5, 6]. From an intervention strategy perspective, there are important differences between the Delta and Omicron variants. While Omicron spreads easier than Delta (even among the vaccinated), it is less likely to cause severe illness [7], especially in vaccinated and boosted populations [8]. Further, while this new period of the pandemic is marked by wide availability of the vaccine, it is also marked by vaccine hesitancy in certain groups [9], and, once more, by the scarcity of the testing resources [10]. As a result, there is an ongoing need to utilize the limited testing resources in the most effective and efficient way for routine screening of the asymptomatic individuals, who can still transmit the disease [11]. Motivated by these observations, in this paper we build and analyze a compartmental model, to develop optimal *customized screening* strategies for college/university campuses, considering the characteristics of the campus population (e.g., vaccine intake rates, campus size, age distribution), and of the disease (e.g., circulating variant(s) and their transmission and disease characteristics, and vaccine effectiveness).

The literature on screening and vaccination for infectious diseases is vast, and growing, thanks to the pandemic. We refer the interested reader to the many references in [12, 13, 14, 15, 16, 17] for screening related work, in [18, 19, 20, 21, 22, 23, 24, 25, 26, 27, 28] for vaccination related work, and in [29, 30, 31] for screening and vaccination related work; and simply discuss the more recent, Omicron-related works here. For instance, [32] develops a compartmental model to predict outcome metrics related to the Omicron variant under different transmission and severity scenarios, while [33] develops a compartmental model that considers vaccination (including boosters), waning immunity from the vaccine, in order to estimate the reduction in transmission rates, and its effect on daily infections, in response to government policies in Korea under different COVID-19 variants. [34] develops a compartmental model to investigate the impact of vaccination coverage on different outcome metrics under the Omicron variant and a less transmissible variant. In addition, [35] builds a model to calculate the conditions required to achieve herd immunity considering multiple vaccine types and COVID-19 variants. Finally, [36] models and studies the effect of hybrid immunity, school reopening, and the Omicron variant in India. Importantly, neither work models and studies screening strategies, which is the main focus of our paper.

Specifically, we contribute to this stream of literature by building an extended compartmental model to study routine screening strategies, which may be customized based on the vaccination status and/or the demographics of campus residents. To this end, we model each individual’s vaccination status using three categories, unvaccinated, fully vaccinated (hereafter, “vaccinated”), and boosted; and consider faculty/staff (hereafter, “faculty”) versus student groups. This customization allows us to consider a wide range of routine screening strategies, ranging from *universal* (i.e., screening all campus residents with the same frequency), to *partially customized* (i.e., screening only selected vaccination status, with a common frequency), to *fully customized* (i.e., further customizing the screening frequency for each group selected for screening). In addition to campus demographics, our compartmental model also accounts for vaccination coverage (proportion vaccinated/proportion boosted), two circulating variants (Delta and Omicron), and less-than-perfect screening compliance of the campus residents.

We also model imperfect vaccine effectiveness, with effectiveness values that depend on both the variant (Delta versus Omicron) and the vaccination status (vaccinated versus boosted); and a time-dependent disease transmission rate, which varies as the proportion of the infectious population changes over time. Because two variants may be circulating at the same time, vaccine effectiveness, infection transmission rates, and other disease characteristics now become conditional on which variant each individual is exposed to.

These aspects necessitate new modeling approaches in our compartmental model. Using this model, we compare the efficiency and effectiveness of various routine screening strategies based on multiple criteria, including the total or peak number of infections/hospitalizations/deaths. Our model allows each college to conduct a comparative study of various screening strategies, enabling them to develop their optimal strategy based on their specific characteristics, such as campus size, vaccination coverage, and available testing resources, as well as the primary metrics of interest to the college.

## Results

The study setting is a hypothetical college with a population of 24,000 (22,500 students and 2,500 faculty/staff), during an 80-day Spring 2022 academic semester that starts in January 2022. Our SEIR (Susceptible, Exposed, Infectious, Removed) framework simulates COVID-19 infection spread over time in a deterministic manner, considering different levels of vaccine-induced immunity and natural immunity, and a variety of protective and preventative interventions, including routine screening of the asymptomatic population, vaccination, face masking, symptomatic testing, and isolation of the test-positives. Another key aspect of this framework is the modeling that the two variants, Delta and Omicron, can be in circulation simultaneously, with parameter *ω*_0_ representing the proportion (in %) of all COVID-19 infections caused by Omicron, and the remaining 1 − *ω*_0_ representing the proportion caused by Delta. We model that vaccine effectiveness, infection transmission, and disease characteristics are conditional on which variant each individual is exposed to. As the initial conditions, some individuals arrive on campus as fully vaccinated or boosted, or with an undetected, asymptotic SARS-CoV-2 infection (135 students and 9 faculty members, representing 0.6% of each group). Extensive sensitivity analyses are conducted to analyze the impact of variations in key model parameters.

### Screening Strategy

We first focus on routine screening that is either universal, or customizes only the screening population (Table 1), and postpone the discussion of screening frequency customization until the next section. The baseline case considers 75% screening compliance, the base-case transmission scenario, and total vaccination coverage of 82% (64% boosted, 18% vaccinated), with 18% unvaccinated at the start of the semester (Table 2). With regards to variant prevalences, we discuss two important cases that represent the pandemic progression: *ω*_*O*_ = 50%, where both Omicron and Delta variants are in circulation in similar rates, which may represent the U.S. during late December 2021 [37]; and *ω*_*O*_ = 95%, where Omicron takes over as the predominant variant, which was the case in the U.S. starting in early January 2022 [6].

**Table 1.** Description of universal and customized screening strategies.

| Screening Strategy | Policy-maker's Decision |
| --- | --- |
| No screening | • None |
| Universal screening | • The common screening frequency |
| Screening population customization | • Which vaccination categories to screen (unvaccinated, vaccinated, boosted)<br>• The common screening frequency for the screening population |
| Screening population and frequency customization | • Which vaccination categories to screen (unvaccinated, vaccinated, boosted)<br>• A screening frequency for each vaccination category included in screening |
| Full customization | • Which vaccination categories (unvaccinated, vaccinated, boosted) and groups (faculty versus students) to screen<br>• A screening frequency for each vaccination category & group (faculty versus students) included in screening |

**Table 2.** Parameter Values and Ranges for Sensitivity Analysis.

| Model Parameter | Value(s) | Comments/References |
| --- | --- | --- |
| **Disease dynamics** |  |  |
| Proportion of infections due to the Omicron variant ($\omega_0$) | 50%, 95% | Assumption with sensitivity analysis |
| Mean latent time | 3.5 days | Average of Delta (4 days) and Omicron (3 days), based on CDC [55], 2021 |
| Mean time to recovery | 5 days | CDC [56], 2021 |
| **Disease transmission** |  |  |
| Ratio of basic reproduction numbers - Omicron:Delta | 3:1 | Ito et al. [49], 2021, Scientific American [50], 2021 and based on Liu & Rocklöv [51], 2022 |
| **Inputs for basic reproduction number of (variant, group) in the best-, base-, worst-case scenarios, respectively:** |  |  |
| Delta, students | 5, 6, 7 | Assumption with sensitivity analysis, based on [57] |
| Omicron, students | $3 \times \{5, 6, 7\}$ | Assumption with sensitivity analysis |
| Delta, faculty | 2.2, 3.2, 4.2 | Assumption with sensitivity analysis, based on [57] |
| Omicron, faculty | $3 \times \{2.2, 3.2, 4.2\}$ | Assumption with sensitivity analysis |
| Reduction in disease transmission rate under a face mask policy ($\gamma$) | 50% | Zhang et al. [58], 2020 |
| **Disease outcomes** |  |  |
| **Vaccine effectiveness against infection for (variant, vaccination status):** |  |  |
| Delta, vaccinated | 80% | Bruxvoort et al. [59], 2021 |
| Omicron, vaccinated | 33% | BMJ [60], 2021 |
| Delta, boosted | 86.7% | Bruxvoort et al. [59], 2021 |
| Omicron, boosted | 69.4% | Paul [61], 2021 |
| Symptom development rate for infected (all vaccination status) | 30% | Assumption (similar to Paltiel et al. [12], 2020), also based on Poletti et al. [62], 2021 |
| Hospitalization rate for symptomatic and unvaccinated (students/faculty) | 1.4% / 8.4% | CDC [63], 2021, COVID-Net [64], 2021, Rabil et al. [30], 2021 |
| **Vaccine effectiveness against hospitalization for symptomatic (variant, vaccination status):** |  |  |
| Omicron, vaccinated | 70% | BMJ [60], 2021 |
| Delta, vaccinated | 91.7% | Andrews et al. [65], 2022 |
| Omicron, boosted | 93% | Based on Thompson et al. [8], 2022 |
| Delta, boosted | 97.5% | Bruxvoort et al. [59], 2021 |
| Fatality rate for hospitalized and unvaccinated (students/faculty) | 0.05% / 2% | Rabil et al. [30], 2021, Statista [66, 67], 2021 |
| **Screening test characteristics** |  |  |
| Test sensitivity (*sens*) | 80% | Based on Stites & Wilen [68], 2020 Woloshin et al. [69], 2020, Yohe [70], 2020 |
| Test specificity (*spec*) | 98% | Yohe [70], 2020 |
| **Coverage characteristics at the start of the semester** |  |  |
| Proportion of vaccinated | 82% | Based on Nietzel [71], 2021, Mauer [72], 2021 and Mayo Clinic [73], 2022 |
| Proportion of boosted | 38% | Based on Anderson [74], 2022 and CDC [75], 2022 |
|  | 64% | Based on Virginia Tech [76], 2022 and CDC [75], 2022 |
| **Sensitivity analysis** |  |  |
| Parameter | Values |  |
| Screening frequency | 1 day, 2 days, 3 days, 7 days, 14 days |  |
| Proportion of infections due to the Omicron variant ($\omega_0$) | 50%, 95% | |
| Coverage (proportion boosted/vaccinated at the start of the semester) | 64% / 18%, 38% / 44%, 0% / 100%, 100% / 0%, 0% / 0% |  |
| Screening compliance ($\eta$) | 75%, 90%, 100% | |
| Transmission severity (basic reproduction number of Delta for faculty/students) | best-case: 2.2 / 5, base-case 3.2 / 6, worst-case 4.2 / 7 |  |

#### Impact of Variant Breakdown

We study the impact of pandemic progression (i.e., the transition of the dominant variant from Delta to Omicron) on screening performance strategies under 82% total vaccination coverage (64% boosted and 18% vaccinated), and 18% unvaccinated. When both Delta and Omicron are circulating at similar rates (*ω*_*O*_ = 50%, Supplementary Table S4), universal screening every 1/2/14 days results in 1,012/4,058/15,458 total infections, and 31/31/142 peak daily infections, respectively. If the boosted individuals are excluded from screening, 1/2/14-day screening yield 7,967/12,168/16,741 total infections, and 45/81/159 peak daily infections, respectively. The strategy that averts the highest number of infections per test is screening the unvaccinated only every 14 days, with 31.6 infections averted per 1,000 tests over no screening (Fig. 1(a)); this strategy also provides the highest reduction in peak infections per test, reducing the peak by 0.9 infections per 1,000 tests over no screening (Fig. 2(a)).

**Figure 1.**
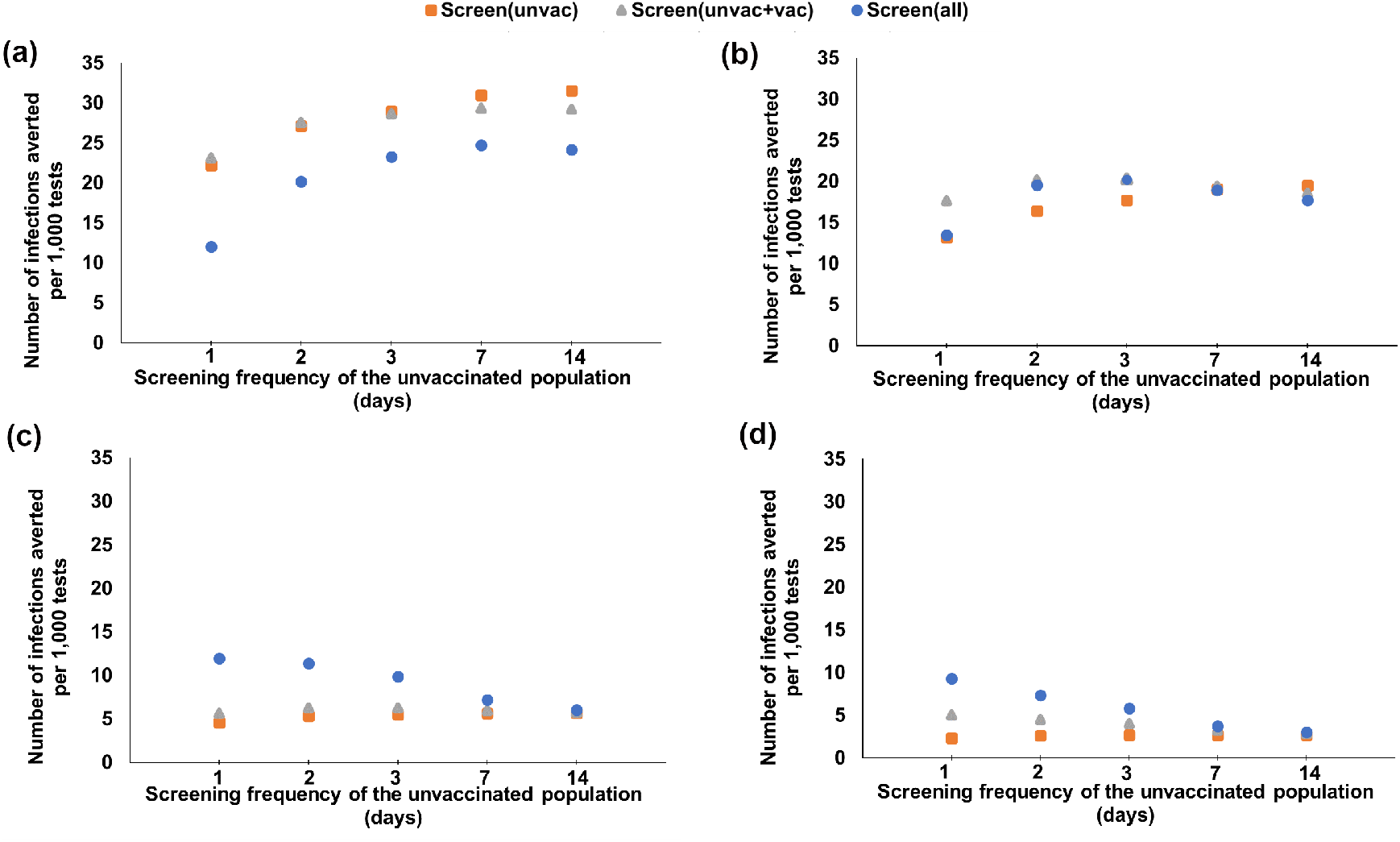
Number of infections averted per 1,000 tests with respect to the screening frequency of the unvaccinated, for various universal and customized screening strategies under various booster and vaccination coverages and Omicron proportions. (a): *ω*_*O*_ = 50%, 64% boosted, 18% vaccinated, 18% unvaccinated, (b): *ω*_*O*_ = 50%, 38% boosted, 44% vaccinated, 18% unvaccinated, (c): *ω*_*O*_ = 95%, 64% boosted, 18% vaccinated, 18% unvaccinated and (d): *ω*_*O*_ = 95%, 38% boosted, 44% vaccinated, 18% unvaccinated

**Figure 2.**
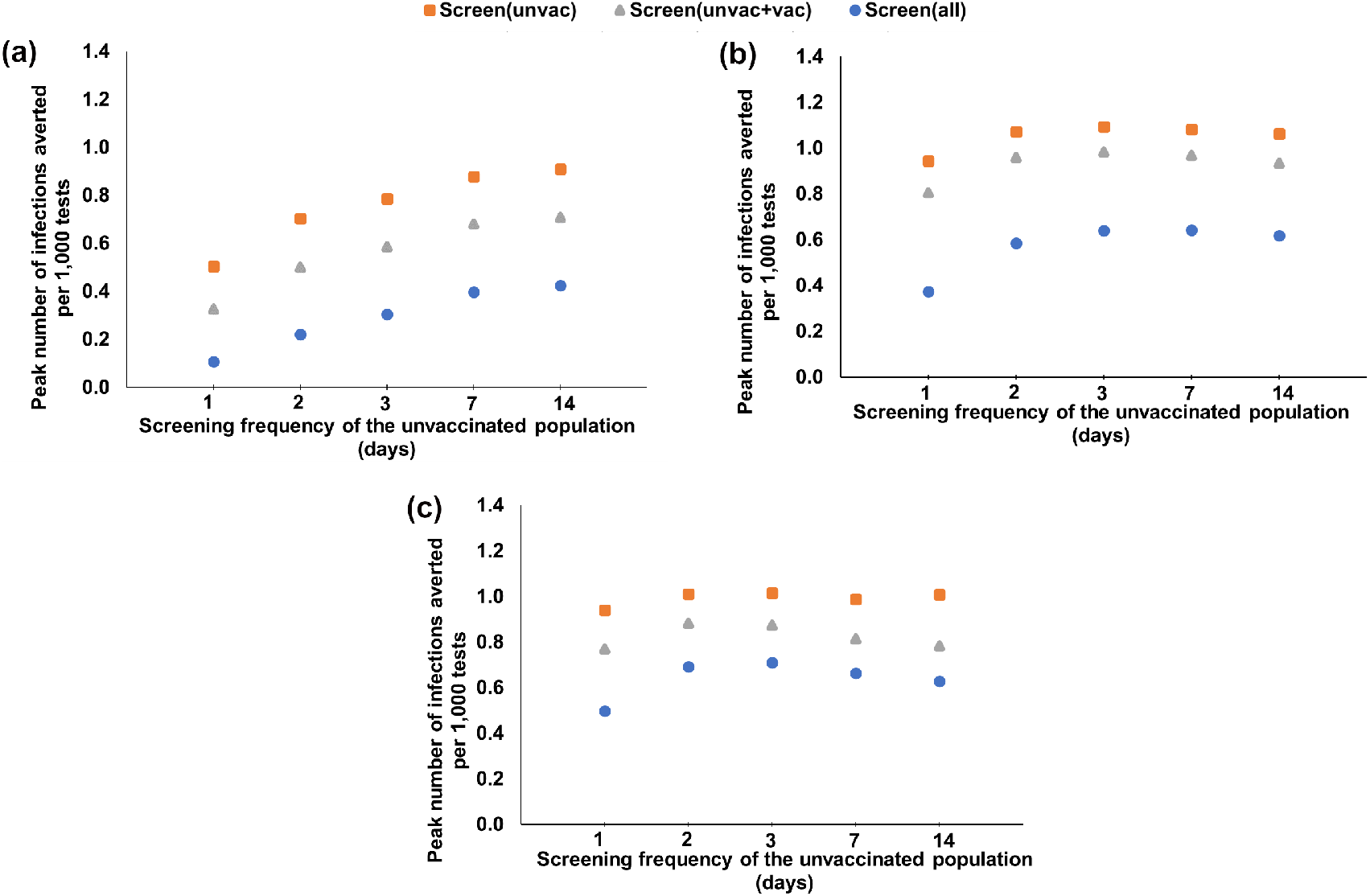
Peak number of infections averted per 1,000 tests with respect to the screening frequency of the unvaccinated, for various universal and customized screening strategies under various booster and vaccination coverages and Omicron proportions. (a): *ω*_*O*_ = 50%, 64% boosted, 18% vaccinated, 18% unvaccinated, (b): *ω*_*O*_ = 95%, 64% boosted, 18% vaccinated, 18% unvaccinated and (c): *ω*_*O*_ = 95%, 38% boosted, 44% vaccinated, 18% unvaccinated

Under *ω*_*O*_ = 50%, the strategy that averts the highest number of infections per test depends on the booster coverage. For the 64% boosted and 18% vaccinated case, if the screening frequency of the unvaccinated is set to every 1/2/3/7/14 days, the strategy that averts the highest number of infections per test is screening the unvaccinated and vaccinated every 1/2 days, and screening the unvaccinated only every 3/7/14 days, averting 23.2/27.5/29/31/31.6 infections per 1,000 tests (Fig. 1(a)), respectively, whereas the strategy that averts the highest number of infections per test with 38% boosted and 44% vaccinated is screening the vaccinated and unvaccinated every 1/2/3/7 days and screening the unvaccinated only every 14 days, averting 17.6/20.19/20.23/19.3/19.5 infections per 1,000 tests (Fig. 1(b)), respectively.

When Omicron is the predominant variant (*ω*_*O*_ = 95%, Supplementary Table S5), universal screening every 1/2/14 days results in 8,568/17,327/22,512 total infections, and 61/222/465 peak daily infections, respectively. When screening excludes boosted individuals, 1/2/14-day screening yields 21,578/22,111/22,820 total infections, and 312/380/487 peak daily infections, respectively. Under this scenario, daily universal screening averts the highest number of infections per test (11.9 per 1,000 tests, Fig. 1(c)), whereas screening the unvaccinated every 3 days reduces the peak infections per test the most (by 1.09 infections per 1,000 tests, Fig. 2(b)).

Under *ω*_*O*_ = 95%, if the unvaccinated is screened every 1/2/3/7/14 days, the strategy that averts the highest number of infections per test is 1/2/3/7/14 day universal screening for both the 64% boosted and 18% vaccinated, or the 38% boosted and 44% vaccinated cases, averting 11.9/9.9/7.2/6 and 7.4/5.8/3.8/3 infections per 1,000 tests (Fig. 1(c) and Fig. 1(d)), respectively.

#### Impact of Vaccination Status

To study the impact of the proportion boosted on screening performance, we first consider that Omicron is the predominant variant (*ω*_*O*_ = 95%). For a total vaccination coverage of 82% (38% boosted, 44% vaccinated, hence 18% unvaccinated), universal screening every 1/2/14 days results in 13,803/20,241/23,221 total infections, and 143/357/612 peak daily infections, respectively (Supplementary Table S5), compared to 8,568/17,327/22,512 total, and 61/222/465 peak daily infections, respectively, when 64% is boosted. Daily universal screening averts the highest number of infections per test (9.3 infections per 1,000 tests, Fig. 1(d), whereas screening the unvaccinated every 3 days reduces the peak infections per test the most (by 1 infection per 1,000 tests, Fig. 2(c)).

The boosted proportion further impacts hospitalizations (Supplementary Table S5). With 38% boosted, universal screening every 1/2/14 days results in 100/133/145 total hospitalizations, and 12/22/29 peak daily hospitalizations, respectively; compared to 61/102/118 total, and 6/14/21 peak daily hospitalizations, respectively, with 64% boosted.

When both Delta and Omicron are circulating at similar rates (*ω*_*O*_ = 50%, Supplementary Table S4) and 38% is boosted, universal screening every 1/2/14 days results in 1,584/7,478/18,546 infections, and peak daily infections of 36/47/219, respectively. If we exclude boosted individuals from screening, 1/2/14-day screening yields 6,341/12,932/19,096 infections, and peak daily infections of 36/94/229, respectively.

### Customizing the Screening Frequencies

In this section, we study the impact of further customizing the screening frequencies. We first consider that the screening frequency can be customized for each vaccination category that is screened, under 82% vaccination coverage (64% boosted, 18% vaccinated). When *ω*_*O*_ = 50%, screening the unvaccinated every 14 days averts the highest number of infections per test (31.6 per 1,000 tests, Fig. 3(a) and Supplementary Table S9). The same strategy also reduces the peak infections per test the most (by 0.9 infections per 1,000 tests, Fig. 4(a)). When *ω*_*O*_ = 95%, the strategy that averts the highest number of infections per test is daily screening of the unvaccinated and vaccinated, and 2-day screening of the boosted (12.4 infections averted per 1,000 tests, Fig. 3(b)), with a 4% improvement compared to the most effective universal screening strategy (i.e., screening the entire population with the same screening frequency), Fig. 3(b) and Supplementary Table S9; whereas screening the unvaccinated only every 3 days reduces the peak infections per test the most (by 1.09 infections per 1,000 tests, Fig. 4(b)).

**Figure 3.**
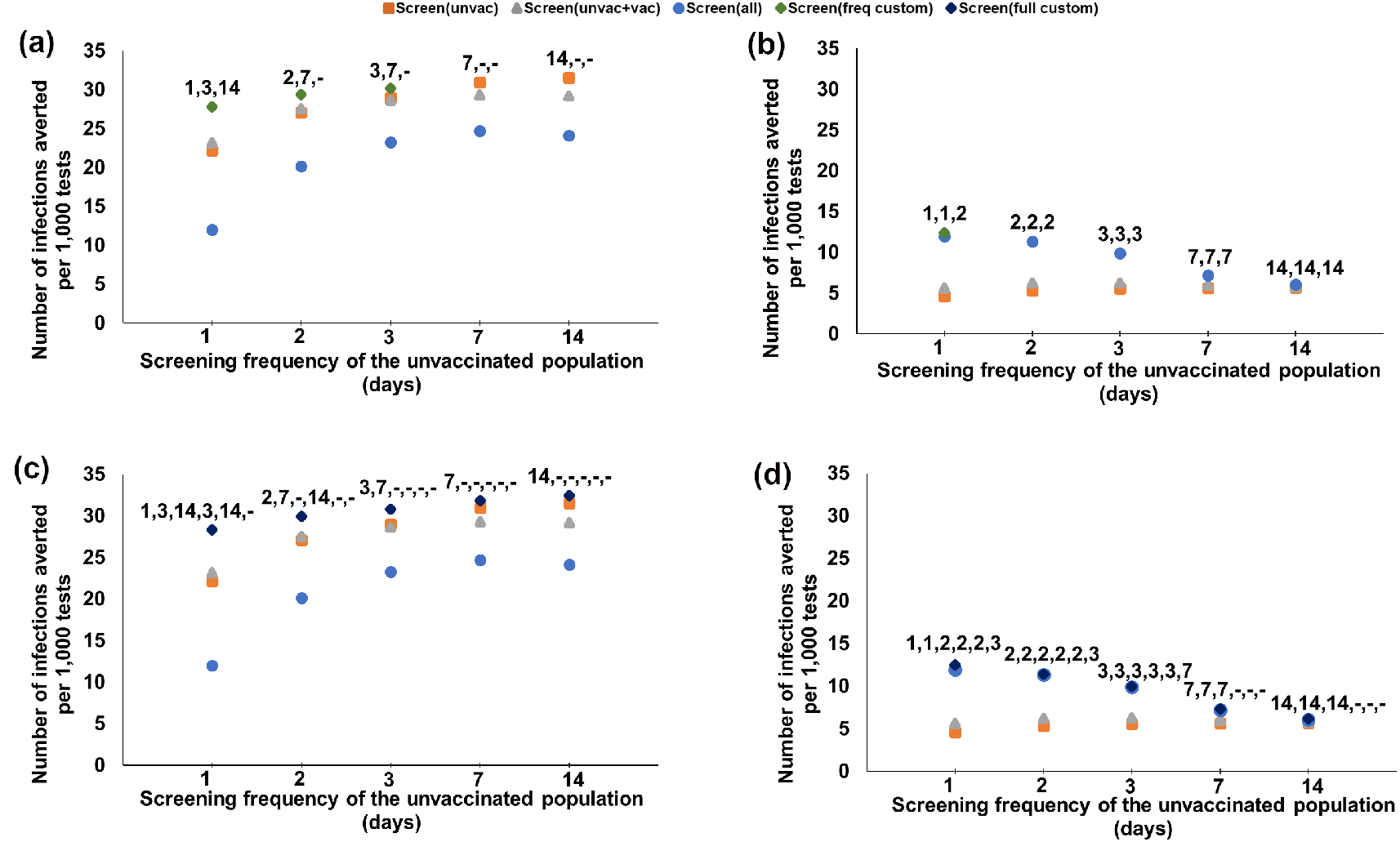
Number of infections averted per 1,000 tests with respect to the screening frequency of the unvaccinated, for various customized screening strategies under 64% boosted, 18% vaccinated, 18% unvaccinated, and various Omicron proportions. (a)-(b): Screening is customized based on vaccination status only; the label represents the screening frequency for unvaccinated, vaccinated, boosted. (c)-(d): Screening is customized based on both vaccination status and faculty versus students; the label represents the screening frequency for unvaccinated students, vaccinated students, boosted students, unvaccinated faculty, vaccinated faculty, boosted faculty. (“-” indicates no screening.)

**Figure 4.**
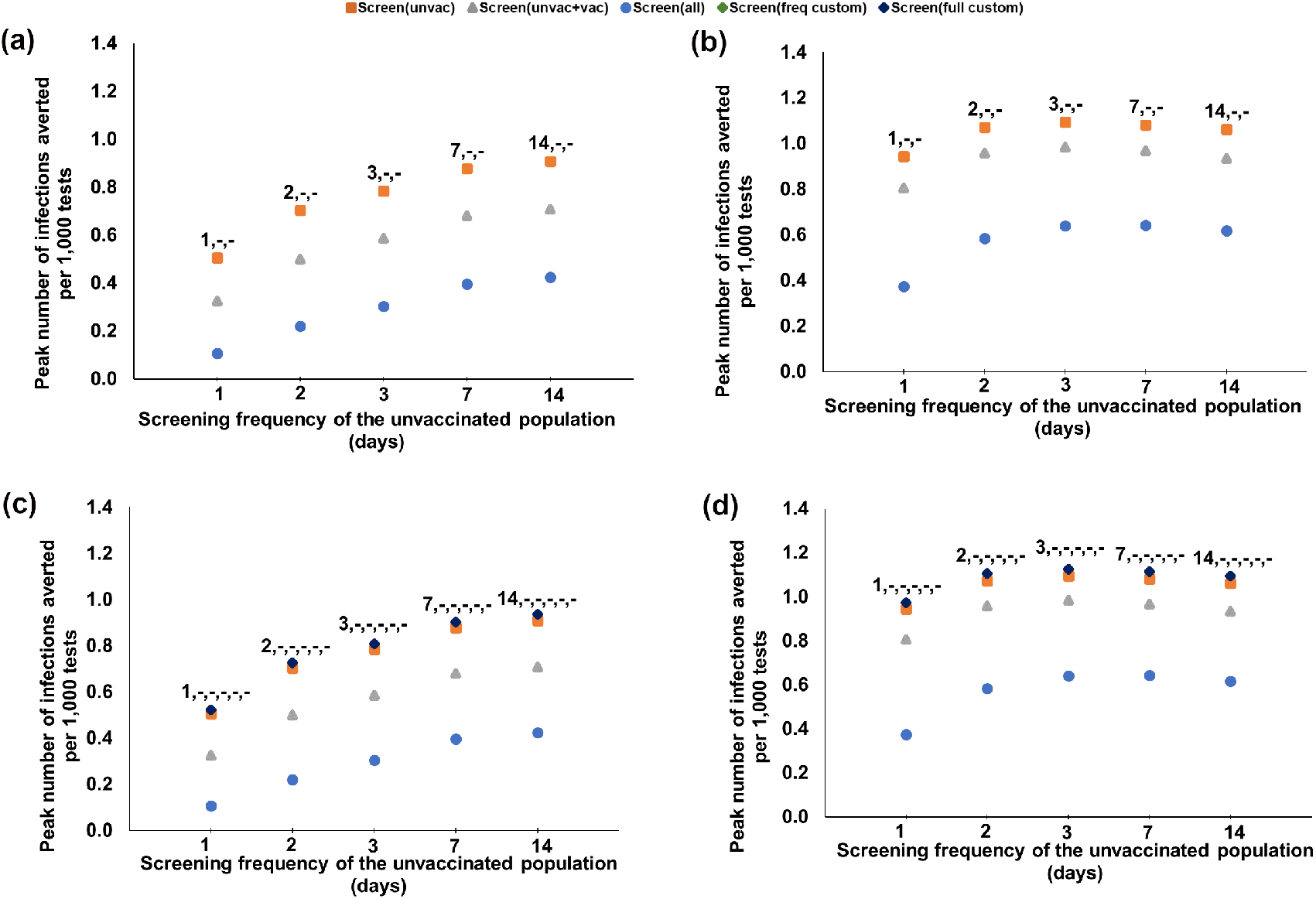
Peak number of infections averted per 1,000 tests with respect to the screening frequency of the unvaccinated, for various customized screening strategies under 64% boosted, 18% vaccinated, 18% unvaccinated, and various Omicron proportions. (a)-(b): Screening is customized based on vaccination status only; the label represents the screening frequency for unvaccinated, vaccinated, boosted. (c)-(d): Screening is customized based on both vaccination status and faculty versus students; the label represents the screening frequency for unvaccinated students, vaccinated students, boosted students, unvaccinated faculty, vaccinated faculty, boosted faculty. (“-” indicates no screening).

We next consider fully customized screening, where the screening frequency can be customized for each group (faculty/students) and each vaccination category that is screened under 82% vaccination coverage (64% boosted, 18% vaccinated). When *ω*_*O*_ = 50%, screening only the unvaccinated students every 14 days averts the highest number of infections per test (32.5 infections averted per 1,000 tests, Fig. 3(c)), with a 2.8% improvement over the most effective strategy that customizes the screening population only. The same strategy also reduces the peak infections per test the, most (by 0.93 infections per 1,000 tests, Fig. 4(c)), providing a 3.3% improvement over the most effective strategy that customizes the screening population only. When *ω*_*O*_ = 95%, the strategy that averts the highest number of infections per test is the daily screening of the unvaccinated and vaccinated students, 2-day screening of the boosted students, and the unvaccinated and vaccinated faculty, and 3-day screening of the boosted faculty (12.5 infections averted per 1,000 tests), with a 5% improvement over the most effective universal screening strategy (Fig. 3(d)); whereas the strategy that provides the highest reduction in peak infections per test is screening only the unvaccinated students every 3 days (by 1.13 infections per 1,000 tests, Fig. 4(d)), providing a 3.7% improvement over the most effective strategy that customizes the screening population only.

### Impact of the Basic Reproduction Number

We study the impact of the basic reproduction number on screening performance by analyzing two additional transmission scenarios (the best- and worst-case, Table 2), considering 64% boosted and 18% vaccinated. When *ω*_*O*_ = 50%, 14-day screening of the unvaccinated averts the highest number of infections per test, with 47.4/31.6/22 infections averted per 1,000 tests under best-/base-/worst-case transmission scenarios, respectively (Supplementary Fig. S2). The same strategy also provides the highest reduction in peak infections per test (with a reduction of 0.8/0.9/1.03 infections per 1,000 tests, Supplementary Fig. S3). When *ω*_*O*_ = 95%, 2/1/1-day universal screening averts the highest number of infections per test, with 15.1/11.9/10.1 infections averted per 1,000 tests under best-/base-/worst-case transmission scenarios, respectively (Supplementary Fig. S2). On the other hand, screening the unvaccinated every 7/3/2-days provides the highest reduction in peak infections per test (by 1.03/1.09/1.13 infections per 1,000 tests, Supplementary Fig. S3).

### Booster Coverage, Vaccine Effectiveness, and Screening Compliance

While screening can be effective for reducing the infection spread, it is uncertain whether aggressive screening, without a high enough booster coverage, would be sufficient for controlling the infection during various phases of the pandemic. We next study the impact of the booster coverage on total infections/hospitalizations under perfect screening compliance for different Omicron proportions and the base-case transmission scenario. If *ω*_*O*_ = 50%, universal daily screening yields 1,036/894/606/473/295 infections, and 10/10/8/7/5 hospitalizations at the end of the semester under booster coverages of 0.2%/20%/50%/75%/82%, respectively, Fig. 5(a), whereas no screening results in 22,445/21,335/19,036/16,919/15,248 infections, and 142/127/103/87/77 hospitalizations at the end of the semester under booster coverages of 0.2%/20%/50%/75%/82%, respectively, Fig. 5(a). If *ω*_*O*_ = 95%, universal daily screening results in 15,965/12,192/5,901/2,382/1,444 total infections, and 127/93/43/20/13 total hospitalizations for booster coverages of 0.2%/20%/50%/75%/82%, respectively, Fig. 5(b), whereas no screening leads to 23,833/23,704/23,338/22,776/22,601 total infections, and 183/164/134/109/101 total hospitalizations under booster coverages of 0.2%/20%/50%/75%/82%, respectively, Fig. 5(b).

**Figure 5.**
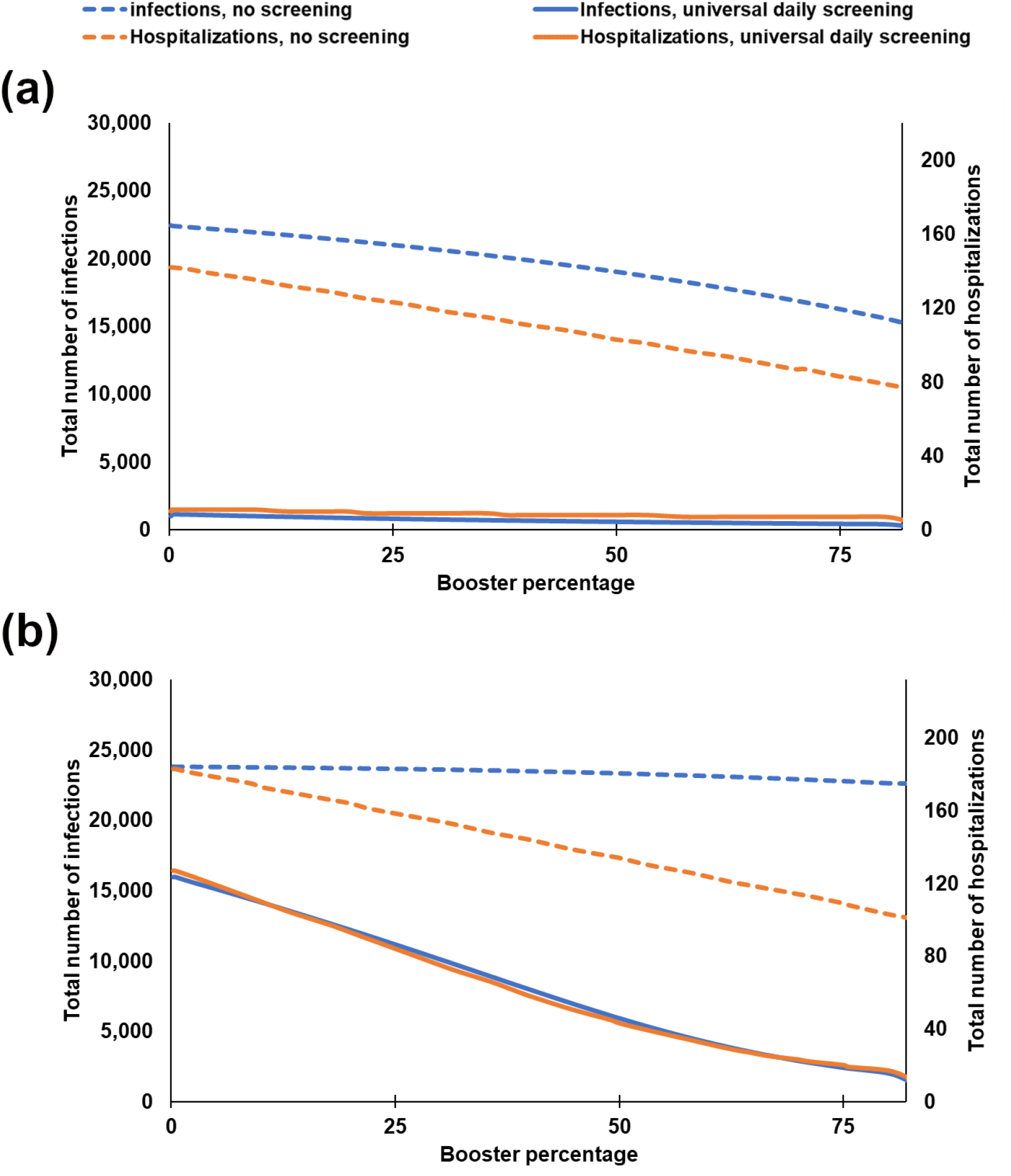
Total number of infections and hospitalizations with respect to the booster coverage under no screening and universal daily screening when 18% is unvaccinated and the remaining is either boosted or vaccinated, and the screening compliance is 100% (*η* = 100%). (a): *ω*_0_ = 50% case, (b): *ω*_0_ = 95% case.

Next, we study the impact of vaccine effectiveness, which is imperfect, time-decaying, and variant-dependent (Table 3), over several outcomes. When *ω*_*O*_ = 95%, even when the entire population is vaccinated (but not boosted) and is screened under perfect compliance, 1/2-day screening leads to 13,755/21,496 infections, and 70/115 hospitalizations under vaccine effectiveness values reported for Omicron (Supplementary Table S7). If the vaccine effectiveness for Omicron were as high as for Delta (Supplementary Table S8), the same level of mitigation efforts would yield 123/327 infections, and 1/2 hospitalization(s). In this scenario, the number of infections and hospitalizations would be 690/4,079/9,068, and 4/17/37, respectively, under 3/7/14-day screening (Supplementary Table S8). The discrepancy between the two scenarios is less striking, but still significant, when Omicron proportion is lower, i.e., *ω*_*O*_ = 50%, or when the entire population is boosted (Supplementary Table S8).

**Table 3.** Weighted Average Computations for the Basic Reproduction Number and Vaccine Effectiveness Values when the Proportion of Infections Caused by Omicron (Delta) is *ω*_0_ (1 − *ω*_0_)

| | Group | Omicron | Delta | Weighted average formula | $\omega_O = 50\%$ case |
| --- | --- | --- | --- | --- | --- |
| Basic reproduction number ($R0$), $k \in \{u, v, b\}$ | **Students** | $3 \times 6 = 18$ | 6 | $R0_{(k,s)}^{\omega_O} = 6 \times (1 - \omega_O) + 18 \times \omega_O$ | $R0_{(k,s)}^{\omega_O=50\%} = 12$ |
| | **Faculty** | $3 \times 3.2 = 9.6$ | 3.2 | $R0_{(k,f)}^{\omega_O} = 3.2 \times (1 - \omega_O) + 9.6 \times \omega_O$ | $R0_{(k,f)}^{\omega_O=50\%} = 6.4$ |
| Vaccine effectiveness against infection ($\epsilon$) | **Vaccinated** | 33% | 80% | $\epsilon_v^{\omega_O} = 80\% \times (1 - \omega_O) + 33\% \times \omega_O$ | $\epsilon_v^{\omega_O=50\%} = 56.5\%$ |
| | **Boosted** | 69.4% | 86.7% | $\epsilon_b^{\omega_O} = 86.7\% \times (1 - \omega_O) + 69.4\% \times \omega_O$ | $\epsilon_b^{\omega_O=50\%} = 78.05\%$ |
| Vaccine effectiveness against hospitalization ($v$) | **Vaccinated** | 70% | 91.7% | $v_v^{\omega_O} = 91.7\% \times (1 - \omega_O) + 70\% \times \omega_O$ | $v_v^{\omega_O=50\%} = 80.85\%$ |
| | **Boosted** | 93% | 97.5% | $v_b^{\omega_O} = 97.5\% \times (1 - \omega_O) + 93\% \times \omega_O$ | $v_b^{\omega_O=50\%} = 95.25\%$ |

Recognizing that different campus populations may exhibit different characteristics in screening compliance, we further investigate the impact of screening compliance. For 82% vaccination coverage (64% boosted, 18% vaccinated) and *ω*_*O*_ = 95%, increasing the compliance rate for universal screening from 75% to 90% reduces the infections from 8,568 to 5,563 for daily screening, and from 17,327 to 15,655 for 2-day screening, Supplementary Table S6.

## Discussion

As new COVID-19 variants emerge, the challenges for effective mitigation evolve. Considering the Spring 2022 academic semester and the co-circulating Delta and Omicron variants, the results from the extended compartmental model in this study suggest that routine screening continues to play a key role in the safe opening and operation of universities. However, allocating the limited screening resources in the most effective manner requires extensive planning, considering the transmission and disease dynamics of the circulating variants, as well as the vaccination coverage, the imperfect, waning, and variant-dependent immunity from vaccination, and the level of natural immunity in the population.

While the benefits of routine screening increase with screening coverage or frequency, a frequent screening of the entire campus population may not be feasible, nor desirable, due to the limited testing resources [38] and the testing fatigue [39]. Thus, designing strategies that yield the highest per-test benefit for key metrics is essential for a safe campus environment, for which an uninterrupted in-person educational experience is important [40]. Because Omicron has relatively short incubation time and high transmissibility, and because many students live on campus, in close-proximity to each other, a high number of infections would threaten in-person teaching. Therefore, we report the total/peak number of infections averted per test as the primary metric. In our setting, with a young and relatively healthy population, hospitalizations/deaths averted per test are low, and quite similar among different screening strategies. Our results demonstrate that the screening strategy that averts the highest number of infections per test depends on the booster coverage and the characteristics of the predominant variant. Comparing universal strategies with those that customize the screening population, our results indicate that, in most cases, universal screening is not the most efficient strategy in terms of infections averted per test when both Delta and Omicron variants are in circulation at similar rates, which may represent the U.S. during late December 2021 [37]. When Omicron is the predominant variant, which has been the case in the U.S. since early January 2022 [6], universal screening does provide the highest per-test reduction in infections. In fact, our findings highlight that the higher the proportion of Omicron and the lower the boosted coverage, the more vaccination status categories need to be screened, and at a higher frequency, in order to maximize the infections averted per test. Several factors, including the higher reproduction number of, and the lower vaccine efficiency against, Omicron, and the waning vaccine-induced immunity against both variants, contribute to this finding. Furthermore, we observe that as the proportion of Omicron (versus Delta) decreases, the screening frequency needed to maximize the efficiency also decreases.

Another key finding is the need for the decision-maker to revisit their mitigation objectives as new variants, with different disease dynamics, emerge. Our results show that when Omicron is the primary variant and screening resources are limited, it might be better to focus on minimizing the peak infections, instead of the total infections, where the latter requires aggressive screening that may not be resource-feasible, or practical, for most universities. We show that screening only unvaccinated individuals (that is, customizing the screening population) is the most efficient strategy in terms of the peak infections averted per test under various Omicron proportions and boosted coverage rates. This finding signifies that when a variant with a higher reproduction number is the dominant strain, allocating the available testing resources to the most vulnerable provides the most efficient response to the pandemic, by “flattening the curve.” We need to emphasize, however, that the most efficient strategy, i.e., that maximizes total or peak infections averted per-test, may not (and in most scenarios will not) minimize the total number of infections. Since expanding the screening coverage and/or frequency always reduces the total number of infections, universities may need to choose their strategy based on sequentially increasing the screening coverage/frequency in the most efficient way, until the expected number of infections is reduced to a tolerable level.

There have been significant discrepancies among U.S. colleges and universities regarding routine screening. While some universities conducted universal screening [41], others customized the screening population based on vaccination status, but still screening them with the same frequency [42, 43], and yet others did switch, at some point, to screening the faculty and students with different frequencies. For instance, at the beginning of the Spring semester, Boston University required the faculty to be routinely screened once a week, and the students twice a week [44]. Stanford, on the other hand, required students to be screened weekly but exempted the faculty from routine screening at some point in the semester [45]. Comparing screening strategies with varying degrees of customization, our findings demonstrate that customizing both the screening population and the frequency based on vaccination status may avert slightly more infections per test over universal screening, or strategies that customize the screening population only, especially when it is feasible to screen the unvaccinated at higher frequencies, and when both Delta and Omicron are in circulation at similar rates. In this case, the most efficient strategy calls for screening the vaccinated and the boosted less frequently than the unvaccinated, and, in cases, not screening the boosted at all. This finding is significant, as it implies that, through customization, a less aggressive strategy (that screens a smaller portion of the population) can provide higher per-test efficiency than universal screening.

Full customization, which considers both the vaccination status and faculty versus student groups further increases the infections averted per test, over customization based on vaccination status alone. While the improvement is small, the main message is again that the decision-maker can achieve higher per-test benefits with less screening. In particular, when full customization is considered, the most efficient strategy (for infections averted per test) recommends the faculty to be screened either less frequently than the students, or not at all. In terms of the peak number of infections averted per test, on the other hand, customizing the screening population already provides a highly efficient strategy, and further customizing the screening frequency does not offer significant benefits: screening the unvaccinated alone remains the dominating strategy. Overall, our results suggest that allowing customization of both the screening population and the frequency based on population characteristics may indeed yield more efficient strategies; equally important is the fact that these customized strategies may also lower the required testing resources during the semester. This is because in many cases efficient customized strategies call for some groups to be either tested with very low frequency, or not at all, which may also reduce the logistical complexity of screening.

As the vaccine-induced immunity in the population wanes over time, or new variants, which are more resistant to the available vaccines, or have lower test efficacy [46], emerge, as was the case at the beginning of 2022, important policy questions arise on whether on-campus screening would be sufficient for infection control, and how these efforts should be adjusted based on the booster coverage at the start of the semester. Our results indicate that when both Delta and Omicron are in circulation at similar rates, aiming for both aggressive screening and high levels of booster coverage may be redundant; screening alone may even be able to compensate for a lack of an appropriate booster coverage. Ideally, integrating moderate levels of booster coverage and screening can provide a highly effective, yet not aggressive, mitigation effort to keep both the number of infections and hospitalizations under control. When Omicron is the primary circulating variant, however, integrating boosters and screening is key for effective mitigation, as none of these efforts would be sufficient, on its own, for controlling the infection, even when implemented aggressively. Accordingly, our results suggest that both adequate booster coverage and routine screening are essential for a safe opening of university campuses, considering the diminishing vaccine effectiveness over time and new vaccine-resistant variant threats. From a practical perspective, integrated screening and booster efforts work especially well towards creating an academic environment that is conducive to in-person learning, because the system is unlikely to be overwhelmed by a large volume of students and faculty missing from the classroom due to an active infection or isolation orders; this can also reduce the potential testing fatigue.

Our sensitivity analyses suggest that if new vaccines are developed with high effectiveness against emerging variants – at levels comparable to the effectiveness of current vaccines for Delta – it may be possible to keep both the infections and hospitalizations under control, even with lenient screening, as long as sufficient levels of vaccination/booster coverage are maintained. Our analysis also indicates that, although higher screening compliance leads to fewer infections, its impact is not substantial, as long as the compliance is kept at a reasonable level (e.g., 75% in our study). Finally, the analysis of the best- and worst-case transmission scenarios indicates that our qualitative results (e.g., the most efficient strategy for given disease and population characteristics, the value of customization and how it depends on model parameters, and which metric to use based on disease and population characteristics) are quite robust to variations in key parameters. On the other hand, the most desirable screening strategy for a given setting depends on the decision-maker’s available testing resources and/or budget, as well as the specific problem parameters. We note that our model can be run in real-time to produce adaptive screening strategies over a semester, as needed (e.g., when large variations in model parameters occur).

## Methods

### Study Design and Parameters

Our SEIR (Susceptible, Exposed, Infectious, Removed) model extends the compartmental framework in [30] in the following ways: We consider a setting where two variants of the virus (Delta and Omicron) might be in circulation simultaneously, and model both vaccine-induced immunity and natural immunity (i.e., acquired based on a prior infection), as well as imperfect vaccine effectiveness that depends on both the variant (Delta versus Omicron) and the vaccination status (vaccinated versus boosted). Due to two circulating variants, vaccine effectiveness, infection transmission, and disease characteristics now become conditional on which variant each individual is exposed to. This setting necessitates the modeling of the heterogeneity in the hypothetical campus population not only in terms of faculty and student groups (indexed by “f, s,” respectively), but also based on vaccination status (unvaccinated, vaccinated, and boosted, indexed by “u, v, b,” respectively), and the presence or absence of natural immunity. The model is a deterministic epidemic model that simulates the transitioning of individuals through different health and vaccination states (compartments), governed by a series of difference equations (based on a cycle of eight hours), and group-dependent and/or vaccination status-dependent transition rates. In particular, each individual transitions through *some* subset of the following health states: exposure to the virus, symptom development or asymptomatic infection, recovery and natural immunity (with or without knowledge of the infection), hospitalization, and death; both disease transmission and disease outcome rates depend on the variant, faculty versus student group, and vaccination status, see the Supplementary information text for details. We consider the following interventions:

- **Isolation and face masking:** All symptomatic subjects, and positive-testing subjects during routine screening immediately go into isolation. Indoor face masking is required for all subjects.
- **Vaccination:** We consider two-dose vaccines (i.e., Pfizer and Moderna, which represent around 96% of the vaccines administered in the U.S. as of March 13, 2022 [47]), and model each subject’s vaccination status through the following categories: unvaccinated, vaccinated (fully vaccinated with a two-dose vaccine prior to August 2021), boosted (fully vaccinated, and boosted in January 2022). We do not consider the population that has received only one dose of a two-dose vaccine. Each subject starts the Spring 2022 academic semester in one of these vaccination categories, and remains in the same vaccination status throughout the semester. We do not consider vaccine mandates, but model the vaccination coverage of the campus population, that is, the proportion boosted/proportion vaccinated. Because the vaccinated individuals (i.e., without a booster) are assumed to have received their second dose more than four months prior to the start of the spring semester, we also model the waning protection from the vaccine, in terms of reduced vaccine effectiveness.
- **Routine screening** excludes subjects who are symptomatic (symptomatic testing is conducted separately), or who are in isolation, at the hospital, or who have tested positive for, and recovered from, the infection (i.e., “recovered and known” subjects). We study routine screening, with the specific strategy dictating the *screening population* (i.e., vaccination categories, or faculty versus student groups included in routine screening) and the *screening frequency* of each vaccination category, or faculty/student group. The screening strategy can be *universal* across all groups and vaccination status categories, or *customized*. We study screening policies, presented below, in increasing level of customization, Table 1.

All screening is conducted via PCR test, the primary test used to detect SARS-CoV-2 [48], and subjects receive their test result after 8 hours, with a positive test result immediately leading to isolation. All false positives are corrected the next day (through additional testing). We assume perfect compliance for all isolation orders and face masking policies, and model imperfect compliance for routine screening.

#### Setting and Parameters

We simulate the infection spread in a hypothetical college of 24,000 (22,500 students and 2,500 faculty members), with 135 students and 9 faculty members (0.6% of each group) having undetected, asymptotic SARS-CoV-2 infection at the outset, and some individuals arriving on campus as vaccinated or boosted. The study period is an 80-day Spring 2022 academic semester that starts in January 2022. Table 2 reports the key parameters, along with corresponding references, and the details are provided in the Supplementary information.

A unique feature of our model is that the two variants, Delta and Omicron, can potentially be in circulation simultaneously, and parameter *ω*_0_ represents the proportion (in %) of all COVID-19 infections caused by Omicron (1 − *ω*_0_ represents the proportion caused by Delta). The basic reproduction number (*R*0) varies with both groups (faculty versus student) and variant (Delta versus Omicron), while vaccine effectiveness against symptom development (*ε*) and hospitalization (*υ*) vary with both vaccination status (vaccinated versus boosted) and variant. To reflect these characteristics in our model, we compute the basic reproduction number per group, and vaccine effectiveness per vaccination status, as *weighted averages* of their respective values for each variant, that is, as a function of *ω*_0_ – Table 3, which provides the key parameters, the weighted average formula, and the computed parameters for the *ω*_0_ = 50% case, considering a 3:1 ratio between the *R*0 values for Omicron and Delta [49, 50, 51]. While these numbers may seem high for the general population, they are more relevant for the college campus setting [52].

#### Outcome Measures of Interest

Total infections, hospitalizations, deaths by group (student versus faculty) and by vaccination status over the 80-day semester; peak daily new infections, peak daily hospitalizations; average number of screening tests per day; number of infections, or peak infections, averted per 1,000 tests compared to no intervention (i.e., no screening), as is common in cost-effectiveness analyses [53]. Further, we use the number of infections averted per test as the primary measure, because Omicron has relatively lower hospitalization and death rates than the Delta variant, especially on a college campus that is mostly populated by a young population with a relatively stronger immunity; and because a high number of infections would pose a threat to an uninterrupted in-person education.

#### Sensitivity Analysis

We conduct sensitivity analyses through varying the values for: the proportion of all COVID-19 infections caused by Omicron (versus Delta), vaccination coverage (proportion boosted/vaccinated), transmission severity, and screening compliance. We study the effectiveness of universal and customized screening strategies (Table 1), obtained by varying the screening frequency(ies) (every 1,2,3,7, 14 days, or no screening) for each vaccination status, Table 2. Different strategies may require different numbers of tests, representing scenarios with different testing capacities or testing kits.

### Statistical Analysis

The compartmental model is coded in C++, and the results are analyzed in Microsoft Excel, through various plots. Our analysis does not involve statistical tests, therefore, we do not report statistical significance levels.

## Code availability

The codes used to generate the results are available at: https://github.com/mjrabil/Screening-for-safe-opening-of-universities, and can be run by a C++ compiler, such as Visual Studio. The instructions for running the codes and generating figures and tables (enclosed in text files) can be found in the README file available at this Github folder. Each code takes at most two minutes to run on a consumer HP laptop having Intel i5 processor and 8GB RAM.

## Conclusion

Routine screening plays a key role in the safe opening of college campuses during a pandemic. Efficiency in screening can only be achieved through customization of strategies based on disease and population characteristics. Our case study of a hypothetical college campus for the Spring 2022 semester, under both Omicron and Delta threats, generates key insights. For example, it indicates that screening the faculty/staff less frequently than the students, or not at all, may avert a higher number of infections per test, compared to less customized strategies, especially when Delta and Omicron variants are in circulation at similar rates. If Omicron is predominant, then a highly effective approach would be to manage the peak infections by routinely screening the unvaccinated population, or even better, the unvaccinated students. Although model parameters are assumed to be deterministic, our sensitivity analyses indicate that the qualitative findings are quite robust to uncertainty; thus we expect our key insights to continue to hold under stochasticity in the values of key parameters. An interesting future extension to our work would be to study routine screening strategies by explicitly modeling the random effects through, for example, a detailed agent-based simulation model, which can model stochasticity at the individual level, e.g. [54].

## Data Availability

All data produced in the present work are contained in the manuscript

## Author contributions statement

M.J.R wrote the first draft of the manuscript, coded the model, and produced the numerical results. S.T. supervised the coding and the visualization of the results. S.T, D.R.B, and E.K.B contributed to the research question, and the conceptualization and supervision of the work. All authors developed the research question and the methodology, analyzed the results, and reviewed and edited the manuscript.

## Data availability statement

All data generated or analysed during this study are included in this published article and its Supplementary Information file as tables and/or figures.

