## Supplementary Information for "Effective screening strategies for safe opening of universities under Omicron and Delta variants of COVID-19"

### Model Description

We developed an extended SEIR (Susceptible, Exposed, Infectious, Removed) framework to model COVID-19 infection spread in a heterogeneous hypothetical population, comprised of faculty and student groups, considering protective and preventative interventions including screening, isolation, masking, and vaccination, when two virus variants are in circulation. The model is a deterministic epidemic model, and tracks the individuals as they transition through different health states (compartments), and the overall flow is governed by a series of difference equations.

In particular, our compartmental model expands that in [9], to consider the following distinct features, see also the flow-chart in Fig. S1:

- We model both vaccine-induced immunity and infection-induced (natural) immunity, that is, an individual can develop protection against a future infection due to either prior vaccination or a prior COVID-19 infection. In particular:
  - We model variant- and dose-dependent vaccine effectiveness. Each individual can be in one of the following compartments based on their vaccination status: "unvaccinated," "vaccinated" (fully vaccinated either with a 1-dose or a 2-dose vaccine prior to August 2021), and "boosted" (fully vaccinated plus received the

booster in January 2022). Because the vaccinated individuals (i.e., without a booster) are assumed to receive their last dose by the start of Fall 2021 semester, we also model waning immunity.

  - We model natural immunity for those individuals who have recovered from a prior infection (through the compartments, recovered & unknown, and recovered & known).

- We model population-based disparities in disease spread, hospitalization, and mortality rates to consider student versus and faculty groups, and their vaccination status. We also consider that the effectiveness of the vaccine depends on whether an individual has received the booster or not, as well as on the circulating virus variant.

- We model variant- and group-dependent disease transmission rates.

In addition, our model preserves the following features from [9]:

- The test result becomes available 8 hours after taking the test. A positive test result indicates either a "false-positive," or "asymptomatic & infected;" and any subject with a positive test result moves to isolation as soon as they receive their test result. No transmission can occur during isolation. All false-positives are corrected the next day. Both the "asymptomatic & infected" and "symptomatic & infected" subjects have an isolation time with a mean of 5 days, after which they move to the "knowingly immune" compartment, unless they are hospitalized. We assume ample isolation capacity and perfect compliance with isolation orders.

- A subject can receive natural immunity through a prior infection; this is modeled through two compartments: The subjects in the "knowingly immune" compartment have gained natural immunity through a prior asymptomatic infection that was detected during routine screening, or a prior symptomatic infection (leading to isolation in each case). The subjects in the "recovered & unknown" compartment have gained natural immunity through a prior asymptomatic infection that was undetected, thus, these subjects cannot be differentiated from "uninfected" individuals, and might get tested.

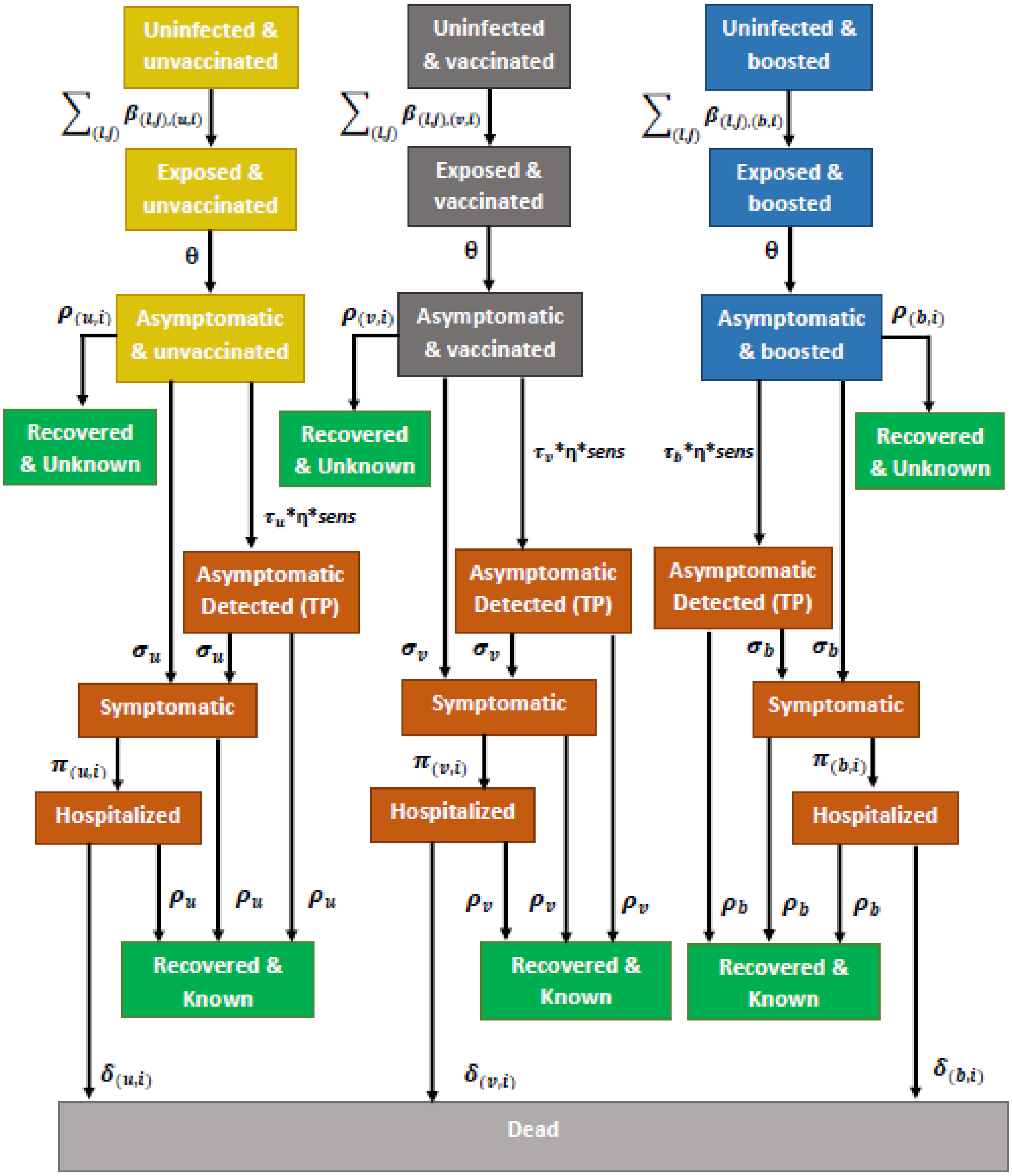


**Supplementary Figure S1:** Flow Diagram of the Extended SEIR Model

**Compartments**

Our model decomposes the population based on group-specific disease and transmission dynamics as well as vaccination status by creating compartments for each population group $i \in \{$(s)tudents, (f)aculty$\}$, and for each vaccination status $k \in \{$(u)nvaccinated, (v)accinated, and (b)oosted$\}$.

We also model that individuals develop (natural) immunity throughout the semester if they have recovered from an infection. Individuals who recover from an infection are assumed to be immune for the rest of the semester. In addition, we assume that the acquired natural immunity and the immunity of the individuals who are boosted do not wane throughout the semester since the it is not long enough (80 days). Accordingly, we have 62 compartments in total, which are categorized into *pools* and described in detail below.

**Transmission pool** contains those individuals who are either susceptible, or currently infected without symptoms and with an unknown infection status (i.e., without a positive test outcome), that is, it contains the following compartments for each group $i$ & vaccination status $k$:

- $U_{k,i}$: Uninfected and susceptible.
- $E_{k,i}$: Exposed, asymptomatic, and non-infectious.
- $A_{k,i}$: Infected and asymptomatic.

**Recovered pool** contains those individuals who have recovered from a prior infection (unknowingly or unknowingly), and hence developed natural immunity, and contains the following compartments for each group $i$ and vaccination status $k$:

- $RU_{k,i}$: Recovered & unknown.
- $RK_{k,i}$: Recovered & known.

**Isolation/hospital pool** contains those individuals who are in isolation or at the hospital. Accordingly, the individuals in this pool cannot transmit the infection, or if they are a false-positive, they cannot get infected. This pool contains the following compartments for each group $i$ & vaccination status $k$:

- $FP_{k,i}$: False-positive with no prior infection (hence in isolation).
- $FPRU_{k,i}$: False-positive, and unknowingly recovered from a prior infection (hence in isolation).
- $TP_{k,i}$ (Asymptomatic Detected): True-positive, infected, and asymptomatic (hence in isolation).
- $S_{k,i}$: Infected and symptomatic (hence in isolation).
- $H_{k,i}$: Hospitalized.

**Removed pool** contains those individuals who have died, that is, for each group $i$, which are contained in the compartment, $D_i$, Dead.

**Screening pool** contains those individuals who are eligible for screening based on the given screening policy.

A summary of the characteristics of each compartment is given in Table S1.

**Supplementary Table S1:** Summary of Compartmental Characteristics

| Compartment | In screening pool? | In transmission pool? | In isolation/ hospital pool? |
|---|---|---|---|
| ($U_{u,i}$)–Uninfected & unvaccinated | Yes | Yes | No |
| ($U_{v,i}$)–Uninfected & vaccinated | Depends on screening policy | Yes | No |
| ($U_{b,i}$)–Uninfected & boosted | Depends on screening policy | Yes | No |
| ($E_{u,i}$)–Exposed & unvaccinated | Yes | Yes | No |
| ($E_{v,i}$)–Exposed & vaccinated | Depends on screening policy | Yes | No |
| ($E_{b,i}$)–Exposed & boosted | Depends on screening policy | Yes | No |
| ($A_{u,i}$)–Asymptomatic & unvaccinated | Yes | Yes | No |
| ($A_{v,i}$)–Asymptomatic & vaccinated | Depends on screening policy | Yes | No |
| ($A_{b,i}$)–Asymptomatic & boosted | Depends on screening policy | Yes | No |
| ($RU_{k,i}$)–Recovered and unknown | Depends on screening policy | No | No |
| ($RK_{k,i}$)–Recovered and known | No | No | No |
| ($FP_{k,i}$)–False-positive with no prior infection | No | No | Yes |
| ($FPRU_{k,i}$)–False-positive, unknowingly recovered from a prior infection | No | No | Yes |
| ($TP_{k,i}$)–True-positive, Asymptomatic detected | No | No | Yes |
| ($S_{k,i}$)–Symptomatic | No | No | Yes |
| ($H_{k,i}$)–Hospitalized | No | No | Yes |
| ($D_i$)–Dead | No | No | No |

### Model Parameters

Subscript $i, j \in \{s, f\}$ denotes the the population group, and subscripts $k, l \in \{u, v, b\}$ denote the vaccination status. When a subscript is omitted, those parameter values apply to all values of the omitted subscript, that is, parameters without the group index $i$ apply to both student and faculty groups, whereas those without any index apply to the entire population (i.e., both groups and all vaccination categories). When needed, we use the superscript"$\omega_O$" to denote probabilistic conditioning, to model the setting where a certain fraction ($\omega_O$) of all COVID-19 infections are caused by the Omicron variant, and the remainder is caused by the Delta variant. We also use the time index ($t$) to indicate that the corresponding parameter varies over time $t$ according to the dynamics of the model.

#### Parameters related to infection prevalence and spread:

$\omega_O$: percentage of all COVID-19 infections caused by the Omicron variant (with the remaining $1 - \omega_O\%$ caused by the Delta variant)

$R0^{\omega_O}_{(l,j)}$: basic reproduction number for subjects in group $j$ & vaccination status $l$, for a given $\omega_O$

$\beta^{\omega_O}_{(l,j),(k,i)}(t)$ rate at which infected subjects in group $j$ & vaccination status $l$ contact and infect subjects in group $i$ & vaccination status $k$, for a given $\omega_O$

$\gamma \in (0,1)$: reduction in disease transmission rate if a face mask policy is implemented

$I_{(k,i)}(t)$: an indicator function, which takes a value of 1 if an exogenous shock takes place in cycle $t$ for group $i$ & vaccination status $k$; and 0 otherwise

$X_{(k,i)}$: number of imported infections per week for subjects in group $i$ & vaccination status $k$ if an exogenous shock takes place in that week

**Parameters related to infection outcomes:**

$\epsilon^{\omega_O}_k$: vaccine effectiveness against infection for subjects with vaccination status $k \in \{v, b\}$, calculated as a weighted average considering an Omicron percentage of $\omega_O$

$\upsilon^{\omega_O}_k$: vaccine effectiveness against hospitalization for subjects with vaccination status $k \in \{v, b\}$, calculated as a weighted average considering an Omicron percentage of $\omega_O$

$\theta$: rate at which exposed subjects become asymptomatic and infectious

$\sigma_k$: rate of symptom onset for infected subjects in vaccination status $k$

$\delta^{\omega_O}_{(k,i)}$: fatality rate for subjects in group $i$ & vaccination status $k$ who are hospitalized

$\pi^{\omega_O}_{(k,i)}$: hospitalization rate for subjects in group $i$ & vaccination status $k$ who are symptomatic

$\rho_k$: recovery rate for infected subjects in vaccination status $k$

**Parameters related to testing:**

$sens$: sensitivity of the screening test

$spec$: specificity of the screening test

$\eta$: screening compliance rate

$\mu$: rate at which subjects with false-positive outcomes return to the uninfected compartment

$\tau_k$: screening rate for subjects with vaccination status $k$

The model uses a cycle time of 8 hours, that is, the number of subjects in each compartment is updated every 8 hours. Screening rate for subjects in each vaccination status, $\tau_k, \forall k$, remains the same throughout the semester. Certain parameter values are varied in the analysis to simulate different strategies or scenarios, e.g., screening frequency, screening compliance rate, etc.

### Governing Equations

The following defines the governing equations for the model depicted in Fig. S1, where $i, j \in \{s, f\}$ and $k, l \in \{u, v, b\}$. Let $Z_{k,i}(t) \equiv U_{k,i}(t) + E_{k,i}(t) + A_{k,i}(t) + RU_{k,i}(t) + RK_{k,i}(t)$.

$$U_{u,i}(t+1) = U_{u,i}(t) \times \left[1 - \sum_l \sum_j \left[\beta^{\omega_O}_{(l,j),(u,i)}(t) \times \frac{A_{l,j}(t)}{Z_{u,i}(t)}\right]\right] - U_{u,i}(t-1) \times \tau_u \times \eta \times (1 - spec)$$
$$+ \mu \times FP_{u,i}(t) - X_{(u,i)} \times I_{(u,i)}(t+1)$$

$$U_{v,i}(t+1) = U_{v,i}(t) \times \left[1 - \sum_l \sum_j \left[\beta^{\omega_O}_{(l,j),(v,i)}(t) \times \frac{A_{l,j}(t)}{Z_{v,i}(t)}\right]\right] - U_{v,i}(t-1) \times \tau_v \times \eta \times (1 - spec)$$
$$+ \mu \times FP_{v,i}(t) - X_{(v,i)} \times I_{(v,i)}(t+1)$$

$$U_{b,i}(t+1) = U_{b,i}(t) \times \left[1 - \sum_l \sum_j \left[\beta^{\omega_O}_{(l,j),(b,i)}(t) \times \frac{A_{l,j}(t)}{Z_{b,i}(t)}\right]\right] - U_{b,i}(t-1) \times \tau_b \times \eta \times (1 - spec)$$
$$+ \mu \times FP_{b,i}(t) - X_{(b,i)} \times I_{(b,i)}(t+1)$$

$$E_{k,i}(t+1) = E_{k,i}(t) \times [1 - \theta] + \sum_l \sum_j \left[\beta^{\omega_O}_{(l,j),(k,i)}(t) \times \frac{U_{k,i}(t) \times A_{l,j}(t)}{Z_{k,i}(t)}\right] + X_{(k,i)} \times I_{(k,i)}(t+1)$$

$$A_{k,i}(t+1) = A_{k,i}(t) \times [1 - \sigma_k - \rho_k] - A_{k,i}(t-1) \times \tau_k \times \eta \times sens + E_{k,i}(t) \times \theta$$

$$FP_{k,i}(t+1) = FP_{k,i}(t) \times [1 - \mu] + U_{k,i}(t-1) \times \tau_k \times \eta \times (1 - spec)$$

$$TP_{k,i}(t+1) = TP_{k,i}(t) \times [1 - \sigma_k - \rho_k] + A_{k,i}(t-1) \times \tau_k \times \eta \times sens$$

$$S_{k,i}(t+1) = S_{k,i}(t) \times [1 - \rho_k - \pi^{\omega_O}_{(k,i)}] + \sigma_k \times [TP_{k,i}(t) + A_{k,i}(t)]$$

$$H_{k,i}(t+1) = H_{k,i}(t) \times [1 - \rho_k - \delta^{\omega_O}_{(k,i)}] + \pi^{\omega_O}_{(k,i)} \times S_{k,i}(t)$$

$$RK_{k,i}(t+1) = RK_{k,i}(t) + \rho_k \times \left[TP_{k,i}(t) + S_{k,i}(t) + H_{k,i}(t)\right]$$

$$RU_{k,i}(t+1) = RU_{k,i}(t) + \rho_k \times A_{k,i}(t) - RU_{k,i}(t-1) \times \tau_k \times \eta \times (1 - spec) + \mu \times FPRU_{k,i}(t)$$

$$D_i(t+1) = D_i(t) + \sum_k \left[\delta^{\omega_O}_{(k,i)} \times H_{k,i}(t)\right]$$

$$FPRU_{k,i}(t+1) = FPRU_{k,i}(t) \times [1-\mu] + RU_{k,i}(t-1) \times \tau_k \times \eta \times (1 - spec)$$

$$N = \sum_k \sum_i \Big[U_{k,i} + E_{k,i} + A_{k,i} + RU_{k,i} + FPRU_{k,i} + S_{k,i} + TP_{k,i} + FP_{k,i} + H_{k,i} + RK_{k,i}\Big] + \sum_i D_i$$

#### Initial Conditions

We assume a 15:1 student to faculty ratio and a medium size college campus, with a total population of 24,000 (22,500 students and 1,500 faculty members). We assume that any subject with some immunity at the beginning of the academic semester has acquired it through vaccination, but model that an individual can acquire natural immunity through an infection during the semester. We consider the following initial conditions, with multiple values for a parameter representing the values considered in sensitivity analysis:

- $A_{k,i}(0) = \begin{cases} 45, & i = s \\ 3, & i = f \end{cases}$
- $U_{u,i}(0) = \begin{cases} \{4,000\ ;\ 7,000\ ;\ 10,000\} - A_{u,i}(0), & i = s \\ \{267\ ;\ 467\ ;\ 667\} - A_{u,i}(0), & i = f \end{cases}$
- $U_{v,i}(0) = \begin{cases} \{4,000\ ;\ 7,000\ ;\ 10,000\} - A_{v,i}(0), & i = s \\ \{267\ ;\ 467\ ;\ 667\} - A_{v,i}(0), & i = f \end{cases}$
- $U_{b,i}(0) = \begin{cases} 22,500 - U_{u,i}(0) - U_{v,i}(0) - A_{b,i}(0) - A_{v,i}(0) - A_{u,i}(0), & i = s \\ 1,500 - U_{u,i}(0) - U_{v,i}(0) - A_{b,i}(0) - A_{v,i}(0) - A_{u,i}(0), & i = f \end{cases}$

All other compartments are initially empty. Accordingly, $N = 22,500 + 1,500 = 24,000$. We note that for a given coverage in students, it is assumed that the coverage in faculty is

15 times less than the coverage in students. This is also assumed to be true within each vaccination status as well, i.e., whenever there is 4K unvaccinated students, it is assumed that there will be 4K/15=267 unvaccinated faculty. In this paper, we consider two options for the coverage in students: (a) 4K unvaccinated, 4K vaccinated, 14.455K boosted, and (b) 4K unvaccinated, 10K vaccinated, 8.455K boosted. This is equivalent to: (a) 82% coverage (64% boosted, 18% vaccinated) and 18% unvaccinated, and (b) 82% coverage (38% boosted, 44% vaccinated) and 18% unvaccinated. We use the latter representation in this paper and omit "18% unvaccinated for simplicity."

Further, in the base case, we use the following values for $X_{k,i}$, the number of imported infections per week on subjects in group $i$ & vaccination status $k$ if an exogenous shock takes place in that week:

- $X_{u,i} = \begin{cases} 15, & i = s \\ 1(= 15/15), & i = f \end{cases}$
- $X_{v,i} = \begin{cases} 10, & i = s \\ 2/3(= 10/15), & i = f \end{cases}$
- $X_{b,i} = \begin{cases} 5, & i = s \\ 1/3(= 5/15), & i = f \end{cases}$

**Estimation of Key Parameters**

We model that the two variants (Delta and Omicron) may be circulating simultaneously, where the parameter $\omega_0$ represents the percentage of all infections caused by the Omicron variant (with $1 - \omega_0$ representing the percentage caused by the Delta variant). Because the reported basic reproduction numbers ($R$) and vaccine effectiveness values ($\epsilon, \upsilon$) differ for the Omicron and Delta variants, we compute the basic reproduction number and vaccine effectiveness values as weighted averages of the respective values for each variant, as a function of $\omega_0$. We consider a 3:1 ratio between the $R$ values for the Omicron and Delta variants [4, 7],

**Supplementary Table S2:** Fatality[1] and Hospitalization Rate Computations for Faculty and Student Groups

| | **Students** | | **Faculty** | |
|---|---|---|---|---|
| | **Hospitalization rate ($H_{(k,s)}$)** | **Fatality rate ($F_{(k,s)}$)** | **Hospitalization rate ($H_{(k,f)}$)** | **Fatality rate ($F_{(k,f)}$)** |
| **For general values of $\omega_O$** | | | | |
| **Unvaccinated** | 1.4% [3, 5, 9] | 0.05% [9, 11, 12] | 8.4% [3, 5, 9] | 2% [9, 11, 12] |
| **Vaccinated** | $1.4\% \times (1 - \upsilon_v^{\omega_O})$ | $0.05\% \times (1 - 1.002 \times \upsilon_v^{\omega_O})$ | $8.4\% \times (1 - \upsilon_v^{\omega_O})$ | $2\% \times (1 - 1.002 \times \upsilon_v^{\omega_O})$ |
| **Boosted** | $1.4\% \times (1 - \upsilon_b^{\omega_O})$ | $0.05\% \times (1 - 1.002 \times \upsilon_b^{\omega_O})$ | $8.4\% \times (1 - \upsilon_b^{\omega_O})$ | $2\% \times (1 - 1.002 \times \upsilon_b^{\omega_O})$ |
| **When $\omega_O = 50\%$** | | | | |
| **Unvaccinated** | 1.4% | 0.05% | 8.4% | 2% |
| **Vaccinated** | 0.2681% | 0.009494% | 1.6086% | 0.3798% |
| **Boosted** | 0.0665% | 0.00228% | 0.399% | 0.09119% |
| $k$ | $\pi_{(k,s)}^{\omega_O}$ | $\delta_{(k,s)}^{\omega_O}$ | $\pi_{(k,f)}^{\omega_O}$ | $\delta_{(k,f)}^{\omega_O}$ |
| **Unvaccinated** | 0.003261 | 0.002469 | 0.0259 | 0.02083 |
| **Vaccinated** | 0.0006007 | 0.002447 | 0.003774 | 0.0206 |
| **Boosted** | 0.000148 | 0.002367 | 0.000898 | 0.01975 |

[1] Vaccine effectiveness against fatality is assumed to be 0.2% higher than vaccine effectiveness against hospitalization

see Table 3 of the Manuscript. Then using these estimates, we find the parameter values reported in Table S2.

In the following, we provide the detailed calculations and references for the computed parameters:

- $\omega_O$, the percentage of all COVID-19 infections caused by the Omicron variant (with the remaining $1 - \omega_0$ caused by the Delta variant). Baseline value of $\omega_O = 75\%$; sensitivity analysis over $\omega_O = \{50\%, 75\%, 95\%\}$.

- $R0_{(l,j)}^{\omega_O}$, basic reproduction number for subjects in group $j$ & vaccination status $l$, for a given $\omega_O$. We assume that for $j \in \{s, f\}$, $R0_{(v,j)}^{\omega_O}$ and $R0_{(b,j)}^{\omega_O}$ are equal to $R0_{(u,j)}^{\omega_O}$ because, once infected, vaccinated subjects are thought to transmit COVID-19 similarly to unvaccinated subjects [10]. Then, we compute $R0_{(u,j)}^{\omega_O}$ as a weighted average of the respective values for the Delta and Omicron variants (see Table 3 of the Manuscript), assuming that Omicron 3 times as infectious as Delta [4, 7]. As of December, 2021, the basic reproduction number for the Delta variant is reported to be between 2 and 8 [6]; and we assume that the basic reproduction number of Delta is 3.2 for the faculty and 6 for the students in the base-case transmission scenario and 2.2/5 and 4.2/7 for the faculty/students in the best- and worst-case scenarios, respectively. Then, when $\omega_O = 50\%$, $R0_{(l,s)}^{\omega_O=50\%} = 12$ and $R0_{(l,f)}^{\omega_O=50\%} = 6.4$, $l \in \{u, v, b\}$ (Table 3 of the Manuscript).

- All vaccine effectiveness values ($\epsilon_k^{\omega_O}, \upsilon_k^{\omega_O}, k \in \{v, b\}$) are computed as weighted averages of the respective values for the Delta and Omicron variants, for a given $\omega_O$ (see Table 3 of the Manuscript). For example, these calculations yield the following numbers for the case of $\omega_0 = 50\%$: $\epsilon_v^{\omega_O=50\%} = 56.5\%$ and $\epsilon_b^{\omega_O=50\%} = 78.05\%$ (i.e., vaccine effectiveness against infection in vaccinated and boosted individuals, respectively), and $\upsilon_v^{\omega_O=50\%} = 80.85\%$ and $\upsilon_b^{\omega_O=50\%} = 95.25\%$ (i.e., vaccine effectiveness against hospitalization in vaccinated and boosted individuals, respectively), see Table 3 of the Manuscript.

- $\beta_{(l,j),(k,i)}^{\omega_O}(t)$, rate at which infected subjects in group $j$ & vaccination status $l$ contact and infect subjects in group $i$ & vaccination status $k$, for a given $\omega_O$. This rate depends on transmission severity, represented in terms of the reproduction number $R_{(l,j),(k,i)}^{\omega_O}$. We model $\beta$ as a function of time ($t$) due to the change in the fraction of susceptibles over time, thus extending the concept of the time-varying reproduction number, described in [1], to a population of subjects with different vaccination status. In particular, in time period $t$, the total number of susceptibles, $N_{su}(t) \equiv \sum_i \sum_k U_{k,i}(t)$, hence we can write, for $i, j \in \{s, f\}, l \in \{u, v, b\}$:

$$\begin{aligned} R_{(l,j),(u,i)}^{\omega_O}(t+1) &= \left[(1-\gamma) \times R0_{(l,j)}^{\omega_O} \times \frac{U_{u,i}(t)}{N_{su}(t)}\right] \\ R_{(l,j),(v,i)}^{\omega_O}(t+1) &= \left[(1-\gamma) \times R0_{(l,j)}^{\omega_O} \times \frac{U_{v,i}(t)}{N_{su}(t)} \times (1-\epsilon_v^{\omega_O})\right] \\ R_{(l,j),(b,i)}^{\omega_O}(t+1) &= \left[(1-\gamma) \times R0_{(l,j)}^{\omega_O} \times \frac{U_{b,i}(t)}{N_{su}(t)} \times (1-\epsilon_b^{\omega_O})\right]. \end{aligned}$$

Then, $\beta_{(l,j),(k,i)}^{\omega_O}(t)$ is the solution to:

$$R_{(l,j),(k,i)}^{\omega_O}(t) = \beta_{(l,j),(k,i)}^{\omega_O}(t)/(\sigma_k + \rho_k) \Rightarrow \beta_{(l,j),(k,i)}^{\omega_O}(t) = R_{(l,j),(k,i)}^{\omega_O}(t) \times (\sigma_k + \rho_k).$$

- $\pi_{(k,i)}^{\omega_O}$, hospitalization rate for subjects in group $i$ & vaccination status $k$ who are symptomatic. These rates are calculated based on hospitalization rates, denoted by $H$ (Table S2), and vaccine effectiveness against hospitalization in vaccinated and boosted subjects $((\upsilon_v^{\omega_O}), (\upsilon_b^{\omega_O})$, Table 3 of the Manuscript). Then, for $i \in \{s, f\}$ and $k \in \{u, v, b\}$, $\pi_{(k,i)}^{\omega_O}$ is the solution to:

$[\sigma_k/(\rho_k + \sigma_k)] \times [\pi_{(k,i)}^{\omega_O}/(\rho_k + \pi_{(k,i)}^{\omega_O})] = H_{(k,i)} \Rightarrow \pi_{(k,i)}^{\omega_O} = \rho_k \times H_{(k,i)}/\big([\sigma_k/(\rho_k + \sigma_k)] - H_{(k,i)}\big)$;

see Table S2 for values of $\pi_{(k,i)}^{\omega_O=50\%}, i \in \{s, f\}, k \in \{u, v, b\}$.

- $\delta_{(k,i)}^{\omega_O}$, fatality rate for subjects in group $i$ & vaccination status $k$ who are hospitalized. These rates are calculated based on fatality rates, denoted by $F$ (Table S2), and vaccine effectiveness against death in vaccinated and boosted subjects, assumed to be 0.2% higher than vaccine effectiveness against hospitalization (i.e., $1.002 \times \epsilon_v^{\omega_O}$ and $1.002 \times \epsilon_b^{\omega_O}$, respectively, see Table 3 of the Manuscript). Then, for $i \in \{s, f\}$ and $k \in \{u, v, b\}$, $\delta_{(k,i)}^{\omega_O}$ is the solution to:

$$[\sigma_k/(\rho_k + \sigma_k)] \times [\delta_{(k,i)}^{\omega_O}/(\rho_k + \delta_{(k,i)}^{\omega_O})] \times [\pi_{(k,i)}/(\rho_k + \pi_{(k,i)})] = F_{(k,i)}$$

$$\Rightarrow \delta_{(k,i)}^{\omega_O} = \rho_k \times F_{(k,i)}/\big([\sigma_k/(\rho_k + \sigma_k)] \times [\pi_{(k,i)}^{\omega_O}/(\rho_k + \pi_{(k,i)}^{\omega_O})] - F_{(k,i)}\big).$$

see Table S2 for values of $\delta_{(k,i)}^{\omega_O=50\%}, i \in \{s, f\}, k \in \{u, v, b\}$.

The following include the parameters that are independent of $\omega_O$:

- $\theta$, rate at which exposed subjects become Asymptomatic & infectious: It is given by $\theta = \frac{1}{33.5} = 0.095$ since the mean latent period is 3.5 days and each day is composed of 3 eight-hour cycles.

- $\sigma_k$, rate of symptom onset in infected individuals with vaccination status $k$: $\sigma_k$ is the solution to, $\sigma_k/(\sigma_k + \rho_k) = 30\%$, where 30% is the probability of developing symptoms after exposure [8], which, in the absence of reliable data, is assumed to be the same for all subjects; and $\rho_k$ is the recovery rate of infected individuals in vaccination status $k$, which, again in the absence of reliable data, is assumed to be the same for all subjects, and is derived from the time to recovery, assumed to be 5 days for all infected individuals [2] (independently of vaccination status and age), where each day is composed of three 8-hour cycles. Then, $\rho_k = 1/(3 \times 5)$, leading to $\sigma_k = 0.0286,\ k \in \{u, v, b\}$.

- $\tau_k$, screening rate for subjects with vaccination status $k$: We have, $\tau_k = 1/(3 \times f_k)$, where $f_k$ is the screening frequency for vaccination status $k$, which can be *daily, every 2 days, every 3 days, every 7 days, or every 14 days.*

- $\gamma$, reduction in disease transmission rate if a face mask policy is implemented. We assume it to be $\gamma = 0.5$ based on [13].

- $\eta$, screening compliance rate: Baseline value of $\eta = 0.75$; with sensitivity analysis over $\eta = \{0.75, 0.90\}$.

Fig. S1 presents a flow diagram of the extended SEIR model. To improve the clarity, Fig. S1 does not include the false-positive compartments, and for those compartments that are defined for both student and faculty groups, only one compartment is shown in the figure. Next, we enclose some results in the following tables. We note that $S/\overline{S}$ denotes screening/no screening, and $u/v/b$ denotes unvaccinated/vaccinated/boosted vaccination status. Thus, $S_u$ and $S_{u,v}$ represent strategies that customize the screening population, $S_{u,v,b}$ represents universal screening, and $\overline{S}$ represents no screening.

**Supplementary Table S3:** Parameter Values and Sensitivity Analysis

| Model Parameter | Value(s) | Input for: |
|---|---|---|
| **Disease related** | | |
| Proportion of infections due to the Omicron variant ($\omega_0$) | 50%, 95% | $R0^{\omega_O}_{(k,i)}$, $\epsilon^{\omega_O}_m$, $\upsilon^{\omega_O}_m$, $H_{(m,i)}$, $F_{(m,i)}$, $R^{\omega_O}_{(l,j),(k,i)}(t)$, $\beta^{\omega_O}_{(l,j),(k,i)}(t)$, $\pi^{\omega_O}_{(m,i)}$, $\delta^{\omega_O}_{(m,i)}$ |
| Mean incubation time | 3.5 days[1] | $\theta$ |
| Time to recovery | 5 days | $\sigma_k$, $\delta^{\omega_O}_{(k,i)}$, $\pi^{\omega_O}_{(k,i)}$ |
| **Infectiousness** | | |
| Infectiousness (basic reproduction number) ratio: Omicron:Delta | 3:1 | $R0^{\omega_O}_{(k,s)}$, $R^{\omega_O}_{(l,s),(k,i)}(t)$, $\beta^{\omega_O}_{(l,s),(k,i)}(t)$ $R0^{\omega_O}_{(k,f)}$, $R^{\omega_O}_{(l,f),(k,i)}(t)$, $\beta^{\omega_O}_{(l,f),(k,i)}(t)$ |
| **Inputs for basic reproduction number of (variant, group):** | | |
| Delta, students | 5, 6, 7 | $R0^{\omega_O}_{(k,s)}$, $R^{\omega_O}_{(l,s),(k,i)}(t)$, $\beta^{\omega_O}_{(l,s),(k,i)}(t)$ |
| Omicron, students | $3 \times \{5, 6, 7\}$ | $R0^{\omega_O}_{(k,s)}$, $R^{\omega_O}_{(l,s),(k,i)}(t)$, $\beta^{\omega_O}_{(l,s),(k,i)}(t)$ |
| Delta, faculty | 2.2, 3.2, 4.2 | $R0^{\omega_O}_{(k,f)}$, $R^{\omega_O}_{(l,f),(k,i)}(t)$, $\beta^{\omega_O}_{(l,f),(k,i)}(t)$ |
| Omicron, faculty | $3 \times \{2.2, 3.2, 4.2\}$ | $R0^{\omega_O}_{(k,f)}$, $R^{\omega_O}_{(l,f),(k,i)}(t)$, $\beta^{\omega_O}_{(l,f),(k,i)}(t)$ |
| Reduction in disease transmission rate under a face mask policy ($\gamma$) | 50% | $R^{\omega_O}_{(l,j),(k,i)}(t)$, $\beta^{\omega_O}_{(l,j),(k,i)}(t)$ |
| **Disease outcomes** | | |
| **Vaccine effectiveness against infection for (variant, vaccination status):** | | |
| Delta, vaccinated | 80% | $\epsilon^{\omega_O}_v$, $R^{\omega_O}_{(l,j),(v,i)}(t)$, $\beta^{\omega_O}_{(l,j),(v,i)}(t)$ |
| Omicron, vaccinated | 33% | $\epsilon^{\omega_O}_v$, $R^{\omega_O}_{(l,j),(v,i)}(t)$, $\beta^{\omega_O}_{(l,j),(v,i)}(t)$ |
| Delta, boosted | 86.7% | $\epsilon^{\omega_O}_b$, $R^{\omega_O}_{(l,j),(b,i)}(t)$, $\beta^{\omega_O}_{(l,j),(b,i)}(t)$ |
| Omicron, boosted | 69.4% | $\epsilon^{\omega_O}_b$, $R^{\omega_O}_{(l,j),(b,i)}(t)$, $\beta^{\omega_O}_{(l,j),(b,i)}(t)$ |
| Symptom development rate for infected (all vaccination status) | 30% | $\sigma_k$, $\delta^{\omega_O}_{(k,i)}$, $\pi^{\omega_O}_{(k,i)}$ |
| Hospitalization rate for symptomatic for unvaccinated (students/faculty) | 1.4% / 8.4% | $H_{(k,i)}$, $\pi^{\omega_O}_{(k,i)}$ |
| **Vaccine effectiveness against hospitalization for symptomatic (variant, vaccination status):** | | |
| Omicron, vaccinated | 70% | $\upsilon^{\omega_O}_v$, $H_{(v,i)}$, $F_{(v,i)}$, $\pi^{\omega_O}_{(v,i)}$, $\delta^{\omega_O}_{(v,i)}$ |
| Delta, vaccinated | 91.7% | $\upsilon^{\omega_O}_v$, $H_{(v,i)}$, $F_{(v,i)}$, $\pi^{\omega_O}_{(v,i)}$, $\delta^{\omega_O}_{(v,i)}$ |
| Omicron, boosted | 93% | $\upsilon^{\omega_O}_b$, $H_{(b,i)}$, $F_{(b,i)}$, $\pi^{\omega_O}_{(b,i)}$, $\delta^{\omega_O}_{(b,i)}$ |
| Delta, boosted | 97.5% | $\upsilon^{\omega_O}_b$, $H_{(b,i)}$, $F_{(b,i)}$, $\pi^{\omega_O}_{(b,i)}$, $\delta^{\omega_O}_{(b,i)}$ |
| Fatality rate for hospitalized for unvaccinated (students/faculty) | 0.05% / 2% | $F_{(k,i)}$, $\delta^{\omega_O}_{(k,i)}$ |
| **Screening test characteristics** | | |
| Test sensitivity (*sens*) | 80% | |
| Test specifity (*spec*) | 98% | |

[1] Average of the 3- and 4-day mean incubation times for Omicron and Delta, respectively

**Supplementary Table S4:** Number of infections (unvaccinated, vaccinated, boosted, total), number of deaths, number of hospitalizations (unvaccinated, vaccinated, boosted, total) over the 80-day semester for various screening strategies, considering that 50% of the infections are caused by Omicron ($\omega_0 = 50\%$), 75% screening compliance ($\eta = 75\%$), and various coverage.

| | | Tests | Infections | | | | | Deaths | Hospitalizations | | | | |
|---|---|---|---|---|---|---|---|---|---|---|---|---|---|
| Strategy | Screening frequency | Average per day | Peak (daily) | Unvaccinated | Vaccinated | Boosted | Total | Total | Peak (daily) | Unvaccinated | Vaccinated | Boosted | Total |
| **82% coverage (64% boosted, 18% vaccinated)** | | | | | | | | | | | | | |
| $\overline{S}$ | N/A | 0 | 180 | 3,729 | 3,356 | 10,522 | 17,606 | 9 | 12 | 72 | 12 | 9 | 93 |
| $S_u$ | every 14d | 179 | 167 | 3,692 | 3,287 | 10,176 | 17,155 | 8 | 11 | 71 | 12 | 9 | 92 |
| | every 7d | 342 | 156 | 3,658 | 3,225 | 9,876 | 16,759 | 8 | 11 | 70 | 12 | 8 | 90 |
| | every 3 d | 739 | 133 | 3,577 | 3,088 | 9,229 | 15,893 | 8 | 10 | 69 | 11 | 8 | 87 |
| | every 2d | 1,070 | 120 | 3,516 | 2,989 | 8,781 | 15,286 | 8 | 9 | 67 | 11 | 7 | 85 |
| | every 1d | 2,065 | 96 | 3,366 | 2,762 | 7,811 | 13,940 | 7 | 7 | 63 | 10 | 6 | 79 |
| $S_{u,v}$ | every 14d | 370 | 159 | 3,656 | 3,227 | 9,859 | 16,741 | 8 | 11 | 70 | 12 | 8 | 90 |
| | every 7d | 720 | 141 | 3,580 | 3,099 | 9,240 | 15,919 | 8 | 10 | 69 | 11 | 8 | 88 |
| | every 3 d | 1,635 | 103 | 3,365 | 2,757 | 7,737 | 13,859 | 7 | 8 | 64 | 10 | 6 | 80 |
| | every 2d | 2,468 | 81 | 3,155 | 2,455 | 6,558 | 12,168 | 7 | 7 | 59 | 8 | 5 | 73 |
| | every 1d | 5,202 | 45 | 2,458 | 1,634 | 3,875 | 7,967 | 5 | 4 | 44 | 5 | 3 | 53 |
| $S_{u,v,b}$ | every 14d | 1,113 | 142 | 3,538 | 3,023 | 8,897 | 15,458 | 8 | 10 | 68 | 11 | 8 | 87 |
| | every 7d | 2,233 | 109 | 3,293 | 2,640 | 7,260 | 13,193 | 7 | 8 | 63 | 9 | 6 | 79 |
| | every 3d | 5,429 | 48 | 2,376 | 1,546 | 3,585 | 7,507 | 5 | 5 | 45 | 6 | 3 | 54 |
| | every 2d | 8,405 | 31 | 1,497 | 826 | 1,735 | 4,058 | 3 | 2 | 29 | 3 | 1 | 34 |
| | every 1d | 17,265 | 31 | 427 | 199 | 386 | 1,012 | 1 | 1 | 11 | 1 | 0 | 12 |
| **82% coverage (38% boosted, 44% vaccinated)** | | | | | | | | | | | | | |
| $\overline{S}$ | N/A | 0 | 263 | 3,873 | 9,210 | 6,959 | 20,042 | 11 | 16 | 75 | 33 | 6 | 114 |
| $S_u$ | every 14d | 173 | 249 | 3,852 | 9,102 | 6,818 | 19,772 | 10 | 16 | 74 | 32 | 6 | 113 |
| | every 7d | 328 | 238 | 3,833 | 9,010 | 6,699 | 19,541 | 10 | 15 | 74 | 32 | 6 | 112 |
| | every 3 d | 691 | 214 | 3,792 | 8,817 | 6,456 | 19,065 | 10 | 14 | 73 | 31 | 6 | 110 |
| | every 2d | 980 | 200 | 3,763 | 8,691 | 6,301 | 18,755 | 10 | 14 | 73 | 31 | 5 | 109 |
| | every 1d | 1,807 | 174 | 3,701 | 8,440 | 6,002 | 18,142 | 10 | 12 | 71 | 30 | 5 | 106 |
| $S_{u,v}$ | every 14d | 637 | 229 | 3,795 | 8,838 | 6,464 | 19,096 | 10 | 15 | 73 | 31 | 6 | 110 |
| | every 7d | 1,238 | 199 | 3,708 | 8,442 | 5,975 | 18,125 | 10 | 14 | 72 | 30 | 5 | 107 |
| | every 3 d | 2,854 | 135 | 3,429 | 7,278 | 4,715 | 15,422 | 9 | 10 | 66 | 25 | 4 | 95 |
| | every 2d | 4,402 | 94 | 3,116 | 6,135 | 3,681 | 12,932 | 8 | 8 | 59 | 21 | 3 | 83 |
| | every 1d | 9,703 | 36 | 1,920 | 2,944 | 1,477 | 6,341 | 4 | 3 | 36 | 9 | 1 | 46 |
| $S_{u,v,b}$ | every 14d | 1,057 | 219 | 3,747 | 8,609 | 6,191 | 18,546 | 10 | 14 | 72 | 31 | 5 | 108 |
| | every 7d | 2,095 | 178 | 3,588 | 7,901 | 5,376 | 16,865 | 9 | 13 | 69 | 28 | 5 | 102 |
| | every 3d | 5,080 | 93 | 2,964 | 5,609 | 3,255 | 11,829 | 7 | 8 | 57 | 19 | 3 | 79 |
| | every 2d | 8,028 | 47 | 2,181 | 3,503 | 1,794 | 7,478 | 5 | 5 | 41 | 12 | 1 | 55 |
| | every 1d | 17,111 | 36 | 573 | 701 | 311 | 1,584 | 1 | 1 | 13 | 3 | 0 | 16 |

**Supplementary Table S5:** Number of infections (unvaccinated, vaccinated, boosted, total), number of deaths, number of hospitalizations (unvaccinated, vaccinated, boosted, total) over the 80-day semester for various screening strategies, considering that 95% of the infections are caused by Omicron ($\omega_0 = 95\%$), 75% screening compliance ($\eta = 75\%$), and various coverage.

| | | Tests | Infections | | | | | Deaths | Hospitalizations | | | | |
|---|---|---|---|---|---|---|---|---|---|---|---|---|---|
| Strategy | Screening frequency | Average per day | Peak (daily) | Unvaccinated | Vaccinated | Boosted | Total | Total | Peak (daily) | Unvaccinated | Vaccinated | Boosted | Total |
| **82% coverage (64% boosted, 18% vaccinated)** | | | | | | | | | | | | | |
| $\overline{S}$ | N/A | 0 | 512 | 4,097 | 4,091 | 14,782 | 22,970 | 11 | 22 | 78 | 23 | 19 | 119 |
| $S_u$ | every 14d | 162 | 498 | 4,091 | 4,083 | 14,722 | 22,896 | 11 | 22 | 78 | 22 | 18 | 119 |
| | every 7d | 300 | 486 | 4,087 | 4,076 | 14,673 | 22,835 | 11 | 22 | 78 | 22 | 18 | 119 |
| | every 3 d | 597 | 459 | 4,076 | 4,060 | 14,568 | 22,705 | 11 | 21 | 78 | 22 | 18 | 119 |
| | every 2d | 812 | 442 | 4,069 | 4,051 | 14,504 | 22,624 | 11 | 21 | 78 | 22 | 18 | 119 |
| | every 1d | 1,361 | 409 | 4,055 | 4,032 | 14,382 | 22,468 | 11 | 20 | 78 | 22 | 18 | 118 |
| $S_{u,v}$ | every 14d | 327 | 487 | 4,085 | 4,075 | 14,659 | 22,820 | 11 | 22 | 78 | 22 | 18 | 119 |
| | every 7d | 608 | 465 | 4,074 | 4,060 | 14,545 | 22,680 | 11 | 21 | 78 | 22 | 18 | 119 |
| | every 3 d | 1,236 | 414 | 4,047 | 4,024 | 14,279 | 22,351 | 11 | 20 | 78 | 22 | 18 | 118 |
| | every 2d | 1,718 | 380 | 4,027 | 3,997 | 14,088 | 22,111 | 11 | 19 | 78 | 22 | 18 | 117 |
| | every 1d | 3,091 | 312 | 3,977 | 3,932 | 13,669 | 21,578 | 11 | 17 | 77 | 22 | 17 | 115 |
| $S_{u,v,b}$ | every 14d | 949 | 465 | 4,063 | 4,041 | 14,408 | 22,512 | 11 | 21 | 78 | 22 | 18 | 118 |
| | every 7d | 1,805 | 419 | 4,019 | 3,976 | 13,936 | 21,931 | 11 | 20 | 78 | 22 | 17 | 117 |
| | every 3d | 4,017 | 306 | 3,843 | 3,722 | 12,228 | 19,794 | 10 | 17 | 74 | 21 | 15 | 110 |
| | every 2d | 6,206 | 222 | 3,608 | 3,396 | 10,323 | 17,327 | 9 | 14 | 70 | 19 | 13 | 102 |
| | every 1d | 15,073 | 61 | 2,334 | 1,902 | 4,331 | 8,568 | 6 | 6 | 45 | 10 | 5 | 61 |
| **82% coverage (38% boosted, 44% vaccinated)** | | | | | | | | | | | | | |
| $\overline{S}$ | N/A | 0 | 658 | 4,133 | 10,482 | 8,828 | 23,443 | 14 | 30 | 78 | 56 | 11 | 146 |
| $S_u$ | every 14d | 160 | 645 | 4,130 | 10,471 | 8,808 | 23,409 | 14 | 30 | 78 | 56 | 11 | 146 |
| | every 7d | 294 | 635 | 4,128 | 10,462 | 8,791 | 23,380 | 14 | 30 | 78 | 56 | 11 | 146 |
| | every 3 d | 578 | 611 | 4,122 | 10,442 | 8,757 | 23,321 | 14 | 29 | 78 | 56 | 11 | 146 |
| | every 2d | 779 | 595 | 4,118 | 10,429 | 8,734 | 23,281 | 14 | 29 | 78 | 56 | 11 | 146 |
| | every 1d | 1,270 | 563 | 4,110 | 10,405 | 8,692 | 23,206 | 14 | 28 | 78 | 56 | 11 | 145 |
| $S_{u,v}$ | every 14d | 565 | 623 | 4,121 | 10,442 | 8,751 | 23,314 | 14 | 29 | 78 | 56 | 11 | 146 |
| | every 7d | 1,049 | 590 | 4,108 | 10,397 | 8,665 | 23,170 | 14 | 29 | 78 | 56 | 11 | 145 |
| | every 3 d | 2,128 | 509 | 4,069 | 10,264 | 8,420 | 22,753 | 13 | 27 | 78 | 55 | 11 | 144 |
| | every 2d | 2,971 | 448 | 4,031 | 10,137 | 8,197 | 22,366 | 13 | 25 | 78 | 55 | 10 | 143 |
| | every 1d | 5,585 | 314 | 3,911 | 9,736 | 7,549 | 21,197 | 13 | 21 | 76 | 52 | 9 | 137 |
| $S_{u,v,b}$ | every 14d | 919 | 612 | 4,113 | 10,411 | 8,697 | 23,221 | 14 | 29 | 78 | 56 | 11 | 145 |
| | every 7d | 1,722 | 566 | 4,087 | 10,314 | 8,525 | 22,925 | 13 | 28 | 78 | 56 | 11 | 145 |
| | every 3d | 3,658 | 450 | 3,978 | 9,915 | 7,848 | 21,741 | 13 | 25 | 77 | 53 | 10 | 140 |
| | every 2d | 5,438 | 357 | 3,830 | 9,384 | 7,026 | 20,241 | 12 | 22 | 74 | 51 | 9 | 133 |
| | every 1d | 12,919 | 143 | 3,023 | 6,766 | 4,014 | 13,803 | 9 | 12 | 59 | 36 | 5 | 100 |

**Supplementary Table S6:** Number of infections (unvaccinated, vaccinated, boosted, total), number of deaths, number of hospitalizations (unvaccinated, vaccinated, boosted, total) over the 80-day semester for various screening strategies, considering that 95% of the infections are caused by Omicron ($\omega_0 = 95\%$), 82% coverage (with 64% boosted), and various screening compliance rates.

| | | Tests | Infections | | | | | Deaths | Hospitalizations | | | | |
|---|---|---|---|---|---|---|---|---|---|---|---|---|---|
| Strategy | Screening frequency | Average per day | Peak (daily) | Unvaccinated | Vaccinated | Boosted | Total | Total | Peak (daily) | Unvaccinated | Vaccinated | Boosted | Total |
| **Screening compliance** $\eta = 75\%$ | | | | | | | | | | | | | |
| $\overline{S}$ | N/A | 0 | 512 | 4,097 | 4,091 | 14,782 | 22,970 | 11 | 22 | 78 | 23 | 19 | 119 |
| $S_u$ | every 14d | 162 | 498 | 4,091 | 4,083 | 14,722 | 22,896 | 11 | 22 | 78 | 22 | 18 | 119 |
| | every 7d | 300 | 486 | 4,087 | 4,076 | 14,673 | 22,835 | 11 | 22 | 78 | 22 | 18 | 119 |
| | every 3 d | 597 | 459 | 4,076 | 4,060 | 14,568 | 22,705 | 11 | 21 | 78 | 22 | 18 | 119 |
| | every 2d | 812 | 442 | 4,069 | 4,051 | 14,504 | 22,624 | 11 | 21 | 78 | 22 | 18 | 119 |
| | every 1d | 1,361 | 409 | 4,055 | 4,032 | 14,382 | 22,468 | 11 | 20 | 78 | 22 | 18 | 118 |
| $S_{u,v}$ | every 14d | 327 | 487 | 4,085 | 4,075 | 14,659 | 22,820 | 11 | 22 | 78 | 22 | 18 | 119 |
| | every 7d | 608 | 465 | 4,074 | 4,060 | 14,545 | 22,680 | 11 | 21 | 78 | 22 | 18 | 119 |
| | every 3 d | 1,236 | 414 | 4,047 | 4,024 | 14,279 | 22,351 | 11 | 20 | 78 | 22 | 18 | 118 |
| | every 2d | 1,718 | 380 | 4,027 | 3,997 | 14,088 | 22,111 | 11 | 19 | 78 | 22 | 18 | 117 |
| | every 1d | 3,091 | 312 | 3,977 | 3,932 | 13,669 | 21,578 | 11 | 17 | 77 | 22 | 17 | 115 |
| $S_{u,v,b}$ | every 14d | 949 | 465 | 4,063 | 4,041 | 14,408 | 22,512 | 11 | 21 | 78 | 22 | 18 | 118 |
| | every 7d | 1,805 | 419 | 4,019 | 3,976 | 13,936 | 21,931 | 11 | 20 | 78 | 22 | 17 | 117 |
| | every 3d | 4,017 | 306 | 3,843 | 3,722 | 12,228 | 19,794 | 10 | 17 | 74 | 21 | 15 | 110 |
| | every 2d | 6,206 | 222 | 3,608 | 3,396 | 10,323 | 17,327 | 9 | 14 | 70 | 19 | 13 | 102 |
| | every 1d | 15,073 | 61 | 2,334 | 1,902 | 4,331 | 8,568 | 6 | 6 | 45 | 10 | 5 | 61 |
| **Screening compliance** $\eta = 90\%$ | | | | | | | | | | | | | |
| $\overline{S}$ | N/A | 0 | 512 | 4,097 | 4,091 | 14,782 | 22,970 | 11 | 22 | 78 | 23 | 19 | 119 |
| $S_u$ | every 14d | 191 | 495 | 4,090 | 4,081 | 14,713 | 22,885 | 11 | 22 | 78 | 22 | 18 | 119 |
| | every 7d | 350 | 481 | 4,085 | 4,073 | 14,654 | 22,812 | 11 | 22 | 78 | 22 | 18 | 119 |
| | every 3 d | 687 | 452 | 4,073 | 4,056 | 14,541 | 22,671 | 11 | 21 | 78 | 22 | 18 | 119 |
| | every 2d | 931 | 433 | 4,066 | 4,046 | 14,472 | 22,584 | 11 | 21 | 78 | 22 | 18 | 119 |
| | every 1d | 1,562 | 400 | 4,050 | 4,027 | 14,350 | 22,427 | 11 | 20 | 78 | 22 | 18 | 118 |
| $S_{u,v}$ | every 14d | 386 | 482 | 4,083 | 4,072 | 14,635 | 22,791 | 11 | 22 | 78 | 22 | 18 | 119 |
| | every 7d | 711 | 456 | 4,070 | 4,055 | 14,501 | 22,626 | 11 | 21 | 78 | 22 | 18 | 119 |
| | every 3 d | 1,433 | 400 | 4,039 | 4,013 | 14,199 | 22,251 | 11 | 20 | 78 | 22 | 18 | 118 |
| | every 2d | 1,996 | 363 | 4,015 | 3,982 | 13,987 | 21,984 | 11 | 19 | 77 | 22 | 18 | 117 |
| | every 1d | 3,649 | 295 | 3,961 | 3,911 | 13,546 | 21,418 | 11 | 17 | 76 | 22 | 17 | 115 |
| $S_{u,v,b}$ | every 14d | 1,125 | 455 | 4,055 | 4,029 | 14,323 | 22,407 | 11 | 21 | 78 | 22 | 18 | 118 |
| | every 7d | 2,134 | 401 | 3,998 | 3,946 | 13,720 | 21,663 | 11 | 20 | 77 | 22 | 17 | 116 |
| | every 3d | 4,850 | 270 | 3,759 | 3,603 | 11,502 | 18,865 | 10 | 16 | 73 | 20 | 14 | 107 |
| | every 2d | 7,714 | 178 | 3,424 | 3,152 | 9,079 | 15,655 | 9 | 12 | 66 | 17 | 11 | 95 |
| | every 1d | 19,090 | 56 | 1,670 | 1,272 | 2,621 | 5,563 | 4 | 3 | 32 | 7 | 3 | 43 |

**Supplementary Table S7:** Total number of infections and total number of hospitalizations over the 80-day semester for various screening strategies, considering various values for the percentage of infections caused by Omicron ($\omega_0 = 0\%, 50\%, 95\%$), 100% screening compliance ($\eta = 100\%$), and extreme cases of coverage.

| | | $\omega_O = 0\%$ | | $\omega_O = 50\%$ | | $\omega_O = 95\%$ | |
|---|---|---|---|---|---|---|---|
| Strategy | Screening frequency | Total number of infections | Total number of hospitalizations | Total number of infections | Total number of hospitalizations | Total number of infections | Total number of hospitalizations |
| **Coverage: 100% boosted** | | | | | | | |
| | N/A | 62 | 0 | 3,823 | 2 | 21,703 | 27 |
| | every 14d | 47 | 0 | 1,290 | 1 | 20,095 | 25 |
| $S_b$ | every 7d | 38 | 0 | 569 | 1 | 17,913 | 21 |
| | every 3 d | 26 | 0 | 183 | 0 | 7,608 | 7 |
| | every 2d | 20 | 0 | 112 | 0 | 1,582 | 2 |
| | every 1d | 12 | 0 | 52 | 0 | 187 | 0 |
| **Coverage: 100% vaccinated** | | | | | | | |
| | N/A | 193 | 0 | 21,748 | 77 | 23,857 | 127 |
| | every 14d | 137 | 0 | 20,191 | 71 | 23,770 | 127 |
| $S_v$ | every 7d | 106 | 0 | 18,094 | 62 | 23,619 | 126 |
| | every 3 d | 66 | 0 | 8,504 | 24 | 22,803 | 122 |
| | every 2d | 50 | 0 | 2,137 | 7 | 21,496 | 115 |
| | every 1d | 29 | 0 | 287 | 1 | 13,755 | 70 |
| **Coverage: 0% (i.e., 100% unvaccinated)** | | | | | | | |
| | N/A | 22,576 | 418 | 23,859 | 441 | 23,919 | 441 |
| | every 14d | 21,545 | 398 | 23,799 | 441 | 23,901 | 441 |
| $S_u$ | every 7d | 20,160 | 371 | 23,689 | 439 | 23,889 | 441 |
| | every 3 d | 14,321 | 244 | 23,059 | 427 | 23,785 | 441 |
| | every 2d | 6,384 | 97 | 22,008 | 408 | 23,547 | 436 |
| | every 1d | 575 | 13 | 15,593 | 285 | 21,508 | 399 |

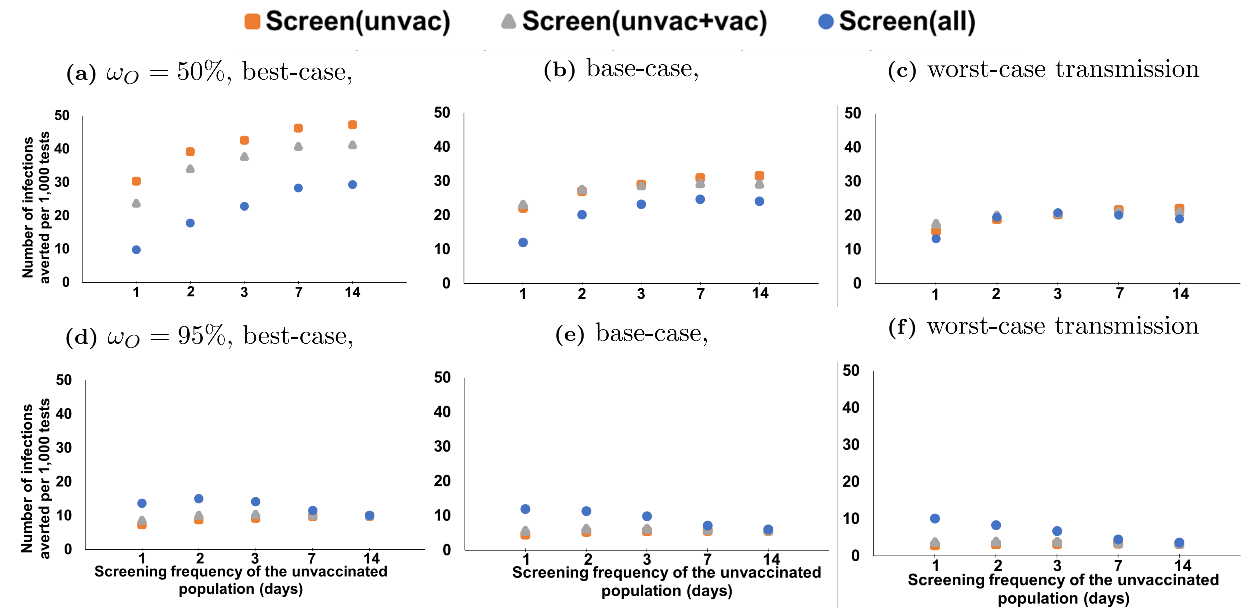


**Supplementary Figure S2:** Number of infections averted per 1,000 tests with respect to the screening frequency of the unvaccinated, for various Omicron proportions and transmission severity scenarios when the vaccination coverage is 64% boosted, 18% vac, 18% unvac (unvac: unvaccinated, vac: vaccinated)

**Supplementary Table S8:** Total number of infections and total number of hospitalizations over the 80-day semester for various screening strategies, considering various values for the percentage of infections caused by Omicron ($\omega_0 = 0\%, 50\%, 95\%$), 100% screening compliance ($\eta = 100\%$), high vaccine effectiveness against infection (as the Delta variant) and extreme cases of coverage.

| | | $\omega_O = 0\%$ | | $\omega_O = 50\%$ | | $\omega_O = 95\%$ | |
|---|---|---|---|---|---|---|---|
| Strategy | Test Frequency | Total number of infections | Total number of hospitalizations | Total number of infections | Total number of hospitalizations | Total number of infections | Total number of hospitalizations |
| **Coverage: 100% boosted** | | | | | | | |
| | N/A | 62 | 0 | 275 | 0 | 1,595 | 2 |
| | every 14d | 47 | 0 | 167 | 0 | 615 | 1 |
| $S_b$ | every 7d | 38 | 0 | 119 | 0 | 327 | 0 |
| | every 3 d | 26 | 0 | 67 | 0 | 134 | 0 |
| | every 2d | 20 | 0 | 48 | 0 | 88 | 0 |
| | every 1d | 12 | 0 | 26 | 0 | 43 | 0 |
| **Coverage: 100% vaccinated** | | | | | | | |
| | N/A | 193 | 0 | 2,477 | 7 | 15,219 | 69 |
| | every 14d | 137 | 0 | 1,001 | 3 | 9,068 | 37 |
| $S_v$ | every 7d | 106 | 0 | 536 | 2 | 4,079 | 17 |
| | every 3 d | 66 | 0 | 218 | 1 | 690 | 4 |
| | every 2d | 50 | 0 | 142 | 1 | 327 | 2 |
| | every 1d | 29 | 0 | 69 | 1 | 123 | 1 |
| **Coverage: 0% (i.e., 100% unvaccinated)** | | | | | | | |
| | N/A | 22,576 | 418 | 23,859 | 441 | 23,919 | 441 |
| | every 14d | 21,545 | 398 | 23,799 | 441 | 23,901 | 441 |
| $S_u$ | every 7d | 20,160 | 371 | 23,689 | 439 | 23,889 | 441 |
| | every 3 d | 14,321 | 244 | 23,059 | 427 | 23,785 | 441 |
| | every 2d | 6,384 | 97 | 22,008 | 408 | 23,547 | 436 |
| | every 1d | 575 | 13 | 15,593 | 285 | 21,508 | 399 |

**Supplementary Table S9:** Number of infections averted per 1,000 tests with respect to the screening frequency of the unvaccinated population under different screening strategies and Omicron proportions for coverage of 82% (64% boosted, 18% vaccinated). (N/A indicates no screening)

| Screening frequency of: | | | | |
|---|---|---|---|---|
| Unvaccinated population | Vaccinated population | Boosted population | Screening strategy, label | Number of infections averted per 1,000 tests |
| $\omega_0 = 50\%$ | | | | |
| every 1d | N/A | N/A | customized screening population, $S_u$ | 22.2 |
| | every 1d | N/A | customized screening population, $S_{u,v}$ | 23.2 |
| | every 1d | every 1d | universal screening, $S_{u,v,b}$ | 12.0 |
| | every 3d | N/A | customized screening population and frequency | 27.8 |
| every 2d | N/A | N/A | customized screening population, $S_u$ | 27.1 |
| | every 2d | N/A | customized screening population, $S_{u,v}$ | 27.5 |
| | every 2d | every 2d | universal screening, $S_{u,v,b}$ | 20.1 |
| | every 7d | N/A | customized screening population and frequency | 29.4 |
| every 3d | N/A | N/A | customized screening population, $S_u$ | 29.0 |
| | every 3d | N/A | customized screening population, $S_{u,v}$ | 28.7 |
| | every 3d | every 3d | universal screening, $S_{u,v,b}$ | 23.3 |
| | every 7d | N/A | customized screening population and frequency | 30.2 |
| every 7d | N/A | N/A | customized screening population, $S_u$ | 31.0 |
| | every 7d | N/A | customized screening population, $S_{u,v}$ | 29.3 |
| | every 7d | every 7d | universal screening, $S_{u,v,b}$ | 24.7 |
| every 14d | N/A | N/A | customized screening population, $S_u$ | 31.6 |
| | every 14d | N/A | customized screening population, $S_{u,v}$ | 29.2 |
| | every 14d | every 14d | universal screening, $S_{u,v,b}$ | 24.1 |
| $\omega_0 = 95\%$ | | | | |
| every 1d | N/A | N/A | customized screening population, $S_u$ | 4.6 |
| | every 1d | N/A | customized screening population, $S_{u,v}$ | 5.6 |
| | every 1d | every 1d | universal screening, $S_{u,v,b}$ | 11.9 |
| | every 1d | every 2d | customized screening population and frequency | 12.4 |
| every 2d | N/A | N/A | customized screening population, $S_u$ | 5.3 |
| | every 2d | N/A | customized screening population, $S_{u,v}$ | 6.3 |
| | every 2d | every 2d | universal screening, $S_{u,v,b}$ | 11.4 |
| every 3d | N/A | N/A | customized screening population, $S_u$ | 5.5 |
| | every 3d | N/A | customized screening population, $S_{u,v}$ | 6.3 |
| | every 3d | every 3d | universal screening, $S_{u,v,b}$ | 9.9 |
| every 7d | N/A | N/A | customized screening population, $S_u$ | 5.6 |
| | every 7d | N/A | customized screening population, $S_{u,v}$ | 6.0 |
| | every 7d | every 7d | universal screening, $S_{u,v,b}$ | 7.2 |
| every 14d | N/A | N/A | customized screening population, $S_u$ | 5.7 |
| | every 14d | N/A | customized screening population, $S_{u,v}$ | 5.8 |
| | every 14d | every 14d | universal screening, $S_{u,v,b}$ | 6.0 |

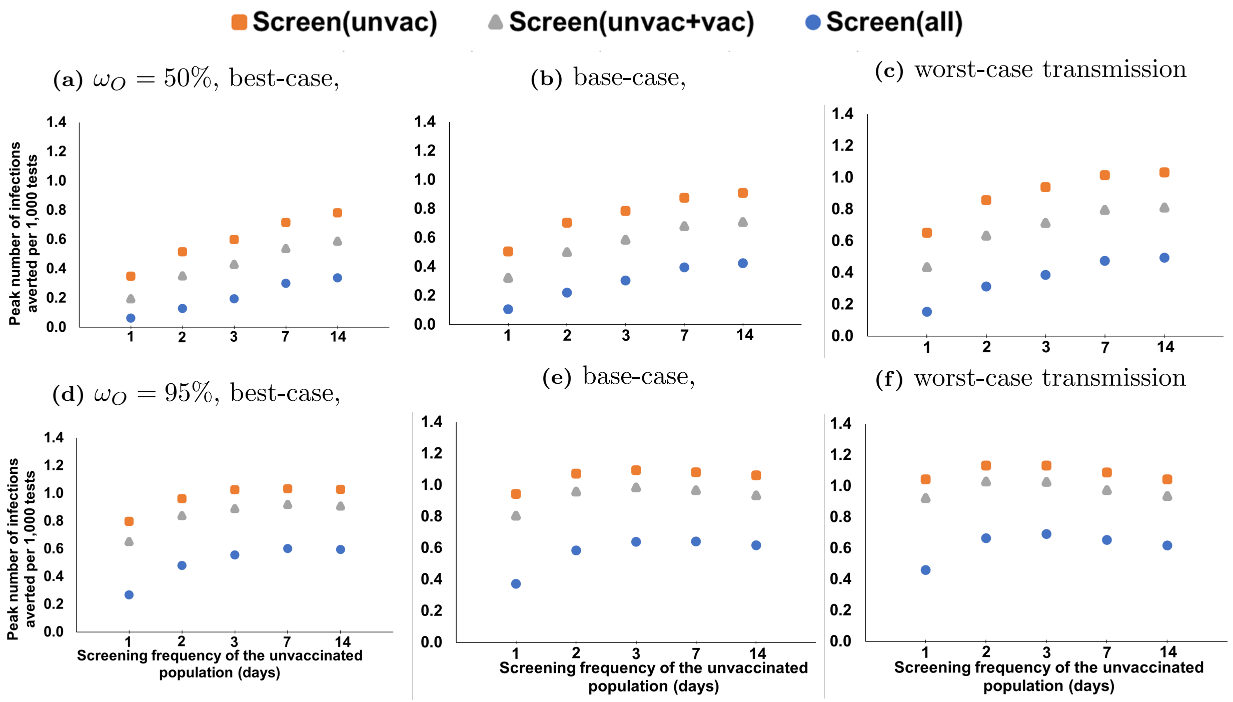


**Supplementary Figure S3:** Peak number of infections averted per 1,000 tests with respect to the screening frequency of the unvaccinated, for various Omicron proportions and transmission severity scenarios when the vaccination coverage is 64% boosted, 18% vac, 18% unvac (unvac: unvaccinated, vac: vaccinated)

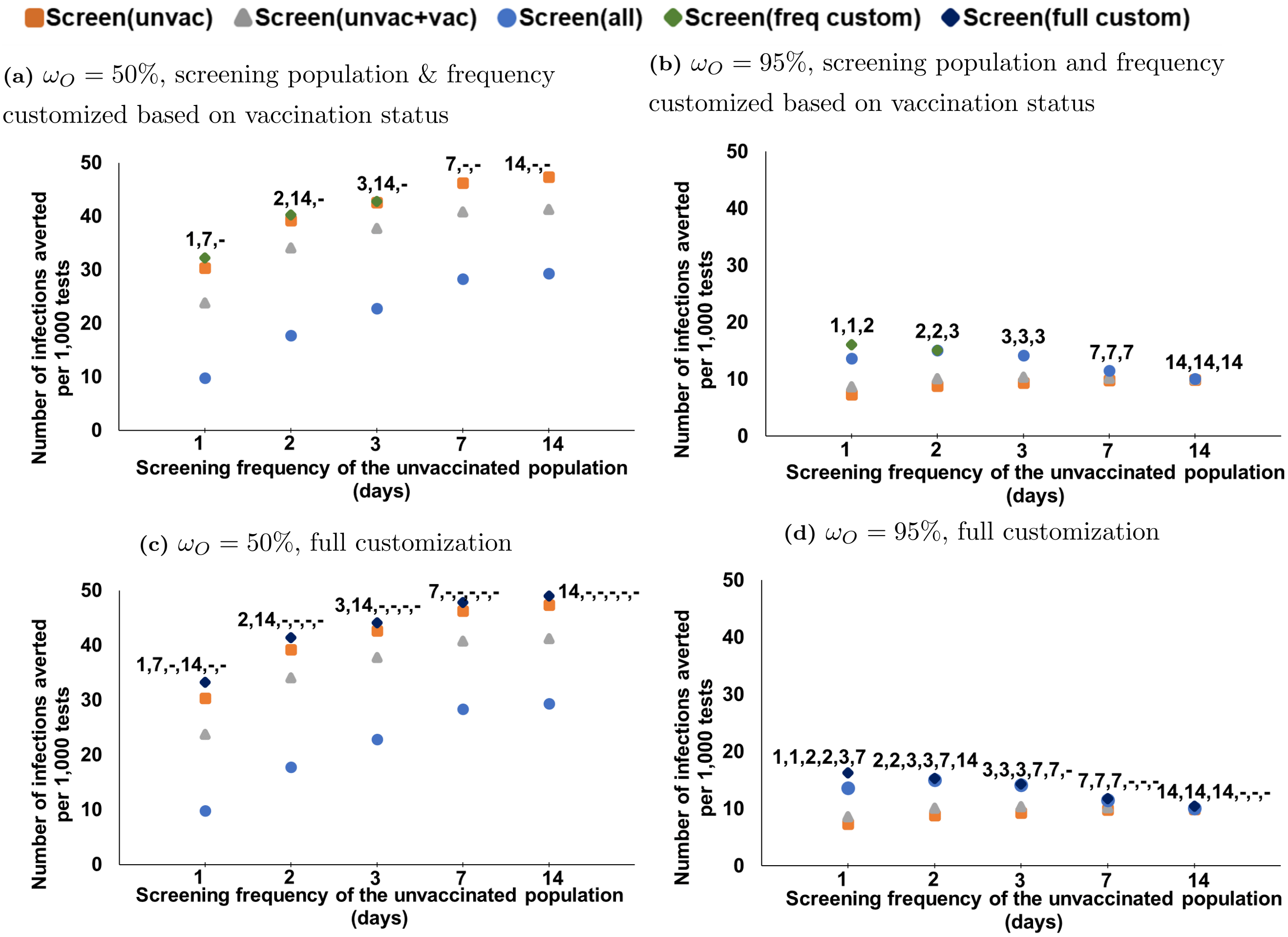


**(a)** $\omega_O = 50\%$, screening population & frequency customized based on vaccination status

**(b)** $\omega_O = 95\%$, screening population and frequency customized based on vaccination status

**(c)** $\omega_O = 50\%$, full customization

**(d)** $\omega_O = 95\%$, full customization

**Supplementary Figure S4:** Number of infections averted per 1,000 tests with respect to the screening frequency of the unvaccinated, for various customized screening strategies under 64% boosted, 18% vaccinated, 18% unvaccinated, best-case transmission and various Omicron proportions. (a)-(b): Screening is customized based on vaccination status only; the label represents the screening frequency for unvaccinated, vaccinated, boosted. (c)-(d): Screening is customized based on both vaccination status and faculty versus students; the label represents the screening frequency for unvaccinated students, vaccinated students, boosted students, unvaccinated faculty, vaccinated faculty, boosted faculty. (“-” indicates no screening.)

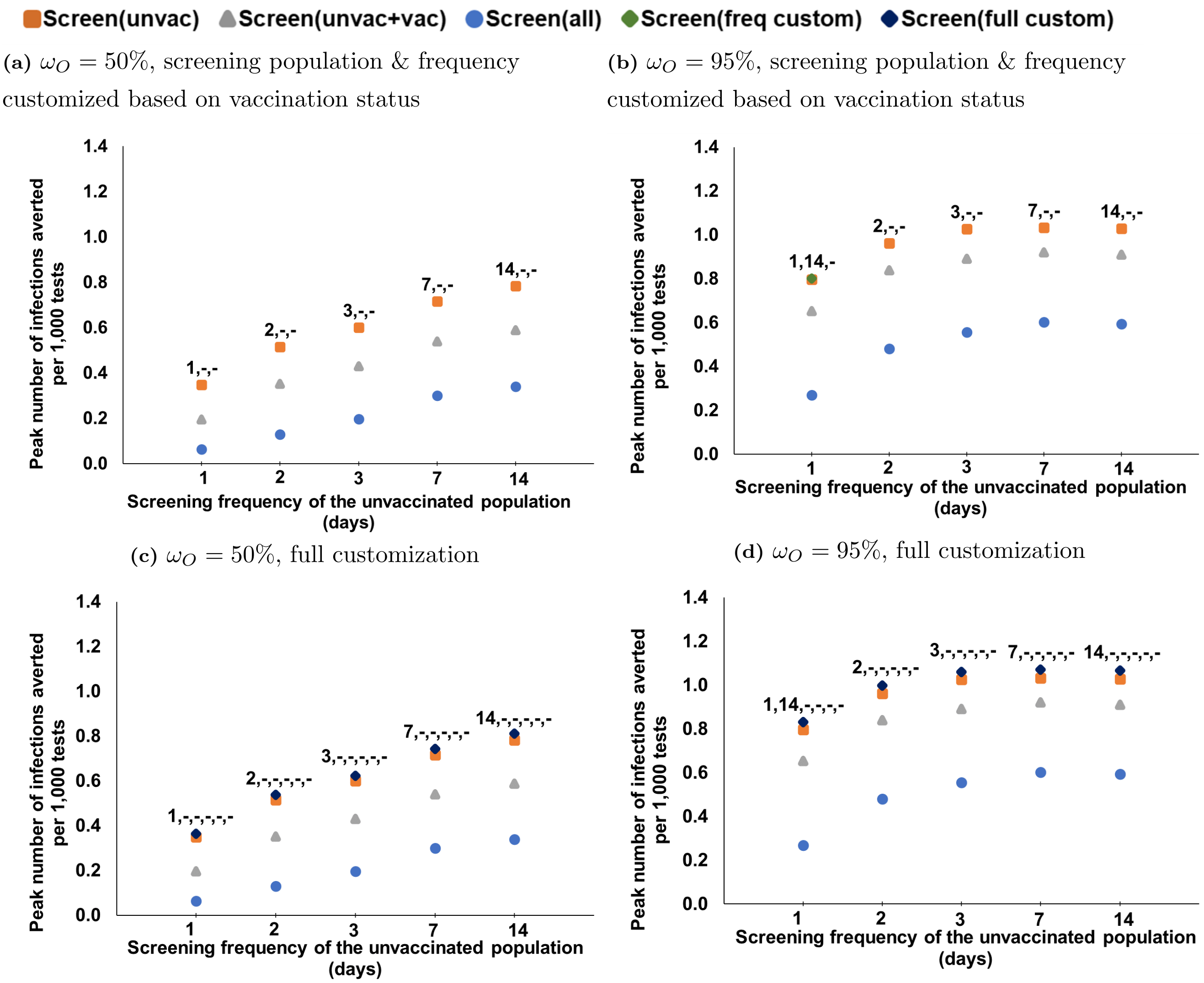


(a) $\omega_O = 50\%$, screening population & frequency customized based on vaccination status

(b) $\omega_O = 95\%$, screening population & frequency customized based on vaccination status

(c) $\omega_O = 50\%$, full customization

(d) $\omega_O = 95\%$, full customization

**Supplementary Figure S5:** Peak number of infections averted per 1,000 tests with respect to the screening frequency of the unvaccinated, for various customized screening strategies under 64% boosted, 18% vaccinated, 18% unvaccinated, best-case transmission and various Omicron proportions. (a)-(b): Screening is customized based on vaccination status only; the label represents the screening frequency for unvaccinated, vaccinated, boosted. (c)-(d): Screening is customized based on both vaccination status and faculty versus students; the label represents the screening frequency for unvaccinated students, vaccinated students, boosted students, unvaccinated faculty, vaccinated faculty, boosted faculty. ("-" indicates no screening).
